# A multicentric, randomized, controlled phase III study of centhaquine (Lyfaquin^®^) as a resuscitative agent in hypovolemic shock patients

**DOI:** 10.1101/2020.07.30.20068114

**Authors:** Anil Gulati, Rajat Choudhuri, Ajay Gupta, Saurabh Singh, S. K. Noushad Ali, Gursaran Kaur Sidhu, Parvez David Haque, Prashant Rahate, Aditya R Bothra, G P Singh, Sanjeev Maheshwari, Deepak Jeswani, Sameer Haveri, Apurva Agarwal, Nilesh Radheshyam Agrawal

**Author notes:** Corresponding author’s complete contact information: Anil Gulati, MD, PhD, Pharmazz, Inc., 50 West 75th Street, Suite 105 Willowbrook, IL 60527, USA, or.

## Abstract

**INTRODUCTION:** Centhaquine (Lyfaquin^®^) showed significant safety and efficacy in preclinical and clinical phase I and II studies.

**METHODS:** A prospective, multicentric, randomized phase III study was conducted in patients with hypovolemic shock having systolic blood pressure (SBP) of ≤90 mm Hg and blood lactate levels of ≥2 mmol/L. Patients were randomized in a 2:1 ratio, 71 patients to the centhaquine group and 34 patients to the control (saline) group. Every patient received standard of care (SOC) and was followed for 28 days. The study drug (normal saline or centhaquine (0.01 mg/kg)) was administered in 100 mL of normal saline infusion over 1 hour. The primary objectives were to determine changes (mean through 48 hours) in SBP, diastolic blood pressure (DBP), blood lactate levels, and base deficit. The secondary objectives included the amount of fluids, blood products, vasopressors administered in the first 48 hours, duration of hospital stay, time in ICU, time on the ventilator support, change in patient’s Acute Respiratory Distress Syndrome (ARDS), Multiple Organ Dysfunction Syndrome (MODS) scores, and the proportion of patients with 28-day all-cause mortality.

**RESULTS:** The demographics of patients and baseline vitals in both groups were comparable. Trauma was the cause of hypovolemic shock in 29.41% of control and 47.06% of centhaquine, gastroenteritis in 44.12% of control, and 29.41% of centhaquine patients. An equal amount of fluids and blood products were administered in both groups during the first 48 hours of resuscitation. A lesser amount of vasopressors was needed in the first 48 hours of resuscitation in the centhaquine group. An increase in SBP from the baseline was consistently higher in the centhaquine group than in the control. A significant increase in pulse pressure in the centhaquine group than the control group suggests improved stroke volume due to centhaquine. The shock index was significantly lower in the centhaquine group than control from 1 hour (p=0.0320) till 4 hours (p=0.0494) of resuscitation. Resuscitation with centhaquine had a significantly greater number of patients with improved blood lactate and the base deficit than the control group. ARDS and MODS improved with centhaquine, and an 8.8% absolute reduction in 28-day all-cause mortality was observed in the centhaquine group.

**CONCLUSION:** Centhaquine is a highly efficacious resuscitative agent for treating hypovolemic shock. The efficacy of centhaquine in distributive shock due to sepsis and COVID-19 is being explored.

**Trial Registration:** Clinical Trials Registry, India; ctri.icmr.org.in, CTRI/2019/01/017196; clinicaltrials.gov, NCT04045327.

**Key Summary Points:**

1. A multicentric, randomized, controlled trial was conducted to evaluate the efficacy of centhaquine in hypovolemic shock patients.
2. One hundred and five patients were randomized 2:1 to receive centhaquine or saline. Centhaquine was administered at a dose of 0.01 mg/kg in 100 mL saline and infused over 1 hour. The control group received 100 mL of saline over a 1-hour infusion.
3. Centhaquine improved blood pressure, shock index, reduced blood lactate levels, and improved base deficit. Acute Respiratory Distress Syndrome (ARDS) and Multiple Organ Dysfunction Syndrome (MODS) score improved with centhaquine.
4. An 8.8% absolute reduction in 28-day all-cause mortality was observed in the centhaquine group. There were no drug-related adverse events in the study.

## INTRODUCTION

Severe blood or fluid loss due to trauma, gastrointestinal bleeding, major surgeries, post-partum, diarrhea, or vomiting can cause hypovolemic shock [1, 2]. About 1.9 million people worldwide die due to hemorrhagic shock every year [3], most dying within the first six hours [4]. Hypovolemic shock’s main features include hypotension, increased blood lactate levels, and base deficit. Hypovolemia decreases cardiac pre-load to a critical level ensuing a dramatic drop in cardiac output that results in low tissue blood perfusion, ultimately leading to multiple organ dysfunction and death. Across the world, patients in intensive care units (ICUs) are resuscitated with fluid therapy to restore blood volume and tissue blood perfusion [5]. Although the goal is to increase the intravascular circulating volume, fluid tends to move out into the extravascular space. An ideal resuscitation fluid should rapidly and effectively increase intravascular volume with sustained effect and minimal third space losses [2].

Numerous attempts have been made to develop an effective resuscitative agent without success. Agents that could decrease metabolic activity to reduce oxygen demand were studied. Histone deacetylase inhibitors [6, 7], hydrogen sulfide and its donor [8, 9], mitochondria-targeted hydrogen sulfide donor AP39 [10], formulation consisting of d-beta-hydroxybutyrate and melatonin [11] and other hibernation based approaches have been tried [12] but none has shown any promise clinically. Hemoglobin-based blood substitutes (oxygen carriers) were developed as resuscitative agents. Diaspirin crosslinked hemoglobin was found effective in animal models of hemorrhagic shock [13, 14], failed in phase III clinical trial [15, 16]. Polymerized hemoglobin, effective in experimental models [17, 18], were not successful clinically [19–21]. Numerous other approaches to developing hemoglobin-based resuscitative agents were not successful [22]. Efforts to develop a pharmacological resuscitative agent have met with failures, and much of the research and development interest in this area has diminished.

Advancement has been limited to damage control resuscitation to restore intravascular volume, prevent dilutional coagulopathy, and preserve tissue oxygenation [23]. An ideal replacement to whole blood is to use blood products in a balanced ratio of 1:1:1 for units of plasma to platelets to red blood cells [24, 25]. Vasopressors are the only pharmacological agents available for resuscitation [26, 27] and are associated with arrhythmias, fluid extravasation, and ischemia [28, 29]. The current standard of care (SOC) is not adequate and is based on resuscitative agents developed more than 5 decades ago, and there is a need for novel resuscitative agents [30].

A substantial amount of blood pooled on the venous side can be returned to the heart and shifted towards the arterial side for better tissue perfusion and oxygenation. Centhaquine is a novel first-in-class resuscitative agent that acts on α_2B_ adrenergic receptors to produce venous constriction and increase venous return to the heart, resulting in increased cardiac output. It also has little action on α_2A_ adrenergic receptors to reduce sympathetic drive and decrease systemic vascular resistance contributing to improved tissue blood perfusion [31]. The mechanism of action of centhaquine makes it an ideal candidate for the treatment of patients with hypovolemic shock. Enhancing tissue blood perfusion is a significant advantage in reducing resuscitation volume and preventing fluid extravasation. Centhaquine has no action on beta-adrenergic receptors, and therefore the possibility of arrhythmias is diminished. The safety and efficacy of centhaquine were evaluated extensively in preclinical models [32–36], healthy volunteers [37, 38] and in patients. Centhaquine was safe and effective in a phase II study with significant improvement in blood lactate levels, base deficit, and blood pressure [39, 40]. Based on these highly encouraging data, a phase III study was undertaken, the results of which are described here.

## METHODS

### Trial Design

This was a prospective, multicenter, randomized, placebo-controlled, double-blinded phase III clinical study of centhaquine in patients with hypovolemic shock receiving the best standard of care (SOC). Because centhaquine was found to be efficacious in a phase II study with statistically significant improvements in blood lactate levels (p=0.0012), base deficit (p= <0.0001) and blood pressure (p=<0.0001) and a trend towards reduced mortality [39, 40], in consultation and agreement with the regulatory authorities, patients were randomized in a 2:1 ratio either to the centhaquine group receiving centhaquine dose of 0.01 mg/kg by IV infusion along with SOC or to the control group receiving SOC plus saline. The study duration for an individual patient was 28 days, including two study visits: visit 1 on day 1 included screening, randomization, baseline measurements, and treatment, while visit 2 was at the end of study (day 28 + 7).

### Regulatory Oversight

The study was conducted in compliance with the Harmonisation of Technical Requirements for Registration of Pharmaceuticals for Human Use Guideline for Good Clinical Practice (ICH-GCP), the Helsinki Declaration, and local regulatory requirements. The study protocol (PMZ-2010/CT-3.1/2018) dated July 16, 2018, was approved by the Drugs Controller General of India (DCGI), Directorate General of Health Services, Ministry of Health & Family Welfare, Government of India (DCGI CT NOC. No.: CT/ND/66/2018). Besides, each institutional ethics committee reviewed and approved the study protocol before initiating patient enrolment. The trial was registered at the Clinical Trials Registry, India (CTRI/2019/01/017196), and the United States National Library of Medicine, ClinicalTrials.gov (NCT04045327). Each site’s ethics committee was informed of any protocol deviation, amendment, subject exclusion or withdrawal, and serious adverse events (SAE). (Details of participating sites are in Supplementary Table 1)

### Patient population

Patients were screened for study eligibility based on the following criteria: 1) patients aged 18 years or older; 2) systolic blood pressure (SBP) of ≤90 mm Hg; 3) blood lactate levels of ≥2 mmol/L that are indicative of hypovolemic shock; 4) patients receiving SOC in a hospital or ICU setting. SOC generally included endotracheal intubation, fluid resuscitation, and vasopressors according to the treatment guidelines in the local hospital setting. Female patients with standing pregnancy were excluded, while patients with postpartum hemorrhage were included. Patients participating in other clinical trials were excluded from the study. Patients with pre-existing systemic diseases such as cancer, chronic kidney failure, liver failure, decompensated heart failure, or AIDS were also excluded.

### Consent

The patients included in this study were in a state of life-threatening shock. For those patients who were not fit to give consent themselves at the time of initiation of treatment, informed consent was taken from their legally authorized representative (LAR). The investigator informed the patient or a LAR in writing and orally details of the study relevant to deciding on participation in the study. The informed consent form included all the elements required as per the ICH-GCP recommendations and schedule Y. Informed consent form in English and regional languages were approved by the respective Ethics Committee and DCGI. The entire consenting process was recorded through an audio-video recording, labeled, and stored at the study site in a secured place. Per the regulatory requirements, medical confidentiality and data protection were ensured, and the investigator stored the signed informed consent forms.

### Randomization and blinding

An Interactive Web Response System (IWRS) containing randomization codes was used to randomize the eligible patient to the treatment groups. The patient and all relevant personnel involved with the study’s conduct and interpretation (including investigator, investigational site personnel, and the sponsor or designee’s staff) were blinded to the study drug’s identity (centhaquine/normal saline) and the randomization codes. The dispensing activity was carried out by unblinded pharmacist independent of the monitoring team. The pharmacist has signed the undertaking of not disclosing the study treatments to the study team. The biostatistician and the unblinded pharmacist were independent of the study team. The final randomization list was held strictly confidential and accessible only to authorized persons until study completion. Emergency unblinding through IWRS was available. As per study protocol, the investigator or his/her designee was permitted to unblind the code when medically needed. For those patients where unblinding was done, the date, time, and the reason for emergency unblinding were recorded in the patient’s medical record. Any adverse event (AE) or SAE that required unblinding the treatment was recorded and reported as specified in the protocol. Treatment unblinding was not done for any of the patients enrolled in this study. Each patient was monitored closely throughout his/her hospitalization and followed until discharge from randomization. Each patient was assessed for safety and efficacy parameters over 28 days from randomization.

### Treatment

At baseline, various demographic data (age, gender, body weight, body mass index), chest X-ray, ECG, vital signs were recorded. Baseline blood tests included hematology, blood lactate, base deficit, serum chemistry, liver, and kidney function tests. The patient’s physical examination, medical history, concomitant illness, concomitant medications, initial Glasgow coma scale (GCS), and Acute Respiratory Distress Syndrome (ARDS) scores were noted. The study drug (1.0 mg of centhaquine citrate in a 10 mL vial) manufactured by Pharmazz India Private Limited at Gufic Biosciences Limited was supplied to the investigators at the participating sites. Patients who met the eligibility criteria were randomized 2:1 to the centhaquine group or control group, respectively. All patients in both groups received the standard of care for hypovolemic shock throughout the study according to local institutional standard practice, including fluid resuscitation with crystalloids/colloids, blood products, and vasopressors. Centhaquine or normal saline was administered intravenously after randomization to hypovolemic shock patients, and all patients continued receiving standard treatment for hypovolemic shock. In the centhaquine group, centhaquine was administered at a dose of 0.01 mg/kg body weight as an intravenous infusion over 1 hour in 100 mL normal saline. The next dose of centhaquine was administered if SBP fell below or remained below 90 mmHg, but not before 4 hours of the previous dose, and the total number of doses did not exceed three per day. Centhaquine administration, if needed, was continued for two days post-randomization. A minimum of 1 dose and a maximum of 6 doses of centhaquine were administered within the first 48 hours post-randomization. An equal volume of normal saline (100 mL) was administered as an intravenous infusion over 1-hour post-randomization in the control group. Specific intravenous treatments and dose selection was based on preclinical proof-of-concept studies conducted in our laboratory [32–36]. The maximum tolerated dose of centhaquine was 0.1 mg/kg as established in the safety and tolerability phase I study [38, 41].

### Data Safety Monitoring Board

A Data and Safety Monitoring Board (DSMB) was convened, and its responsibilities were determined before the study’s initiation. The members included a senior practicing physician with extensive experience in critical care medicine, a biostatistician, and a clinical pharmacologist. The DSMB was independent of the study investigators and the sponsor. The DSMB had access to SAEs and any other AEs that the investigator or the medical monitor considered important. The DSMB reviewed the study data on safety and critical efficacy endpoints at pre-determined intervals.

### Safety Evaluation

All patients who received treatment were included in the safety analysis. Safety was assessed during the treatment period and during the follow-up period post-treatment based on adverse events, physical examination, vital signs (heart rate, SBP and diastolic blood pressure (DBP), body temperature, and respiratory rate), ECG; and clinical laboratory parameters as per protocol. A variety of biochemical tests, serum chemistry tests, hematological variables, coagulation variables, urine output, organ function tests such as kidney and liver function tests were assessed. AEs that occurred or worsened during treatment or post-treatment were recorded. All AEs were coded by systems organ class and preferred term using the latest MedDRA version. All patients were followed up for safety assessment till the end of the study on day 28.

### Efficacy Assessments

This study’s primary objectives were to determine: 1) change in SBP and DBP, 2) change in blood lactate levels, and 3) change in the base deficit. For all these endpoints, changes were mean through 48 hours. The study’s key secondary objectives included the proportion of patients with 28-day all-cause mortality. The amount of fluids, blood products, vasopressors administered in the first 48 hours, duration of hospital stay, time in ICU, time on the ventilator support, patient’s ARDS scores, and multiple organ dysfunction scores (MODS) were recorded.

### Sample Size and Statistical Analysis

The sample size was calculated based on our phase II trial (CTRI/2017/03/008184, NCT04056065) results [39, 40]. In the phase II study, SBP in the centhaquine group was 7.19% higher than the control group at 48 hours (25.39% increase from the baseline in the control group compared to a 39.51% increase from the baseline in the centhaquine group). The statistical power of the study was 80%. Because centhaquine was found to be efficacious in a phase II study with statistically significant improvements in blood lactate levels (p=0.0012), base deficit (p<0.0001) and blood pressure (p<0.0001), and a trend towards reduced mortality, in consultation and agreement with the regulatory authorities, patients were randomized in a 2:1 ratio either to the centhaquine group (SOC + centhaquine) or to the control group (SOC + saline) in the phase III study. The power was set to 90% (beta, 0.1), the enrollment ratio of 2:1, and the significance level (alpha) used was 0.05. To achieve this, we estimated that a sample size of 69 patients (46 in the centhaquine group and 23 in the normal saline group) was enough to achieve a power of 90% when the level of significance alpha was 0.05. In order to increase the power of the study (>90%, beta, 0.05), approximately 84 patients (56 in the centhaquine group and 28 in the control group) were needed to be enrolled. Considering a discontinuation rate of 20%, we planned enrollment of 105 patients (70 in the centhaquine group and 35 in the control group) in this study. The results of the trial are presented as mean ± SEM. Unpaired t-test with Welch’s correction was used to analyze two sets of data with unequal variances. The Unpaired t-test was used to compare the discrete variables between the two sets of data at baseline and follow-ups. Non-parametric analysis was carried out using one-way ANOVA without assuming equal variances, and Tukey’s multiple comparisons test estimated the significance of differences. Group comparison was carried out using a Chi-square test. Baptista-Pike method was used to calculate the odds ratio (OR). A p-value of less than 0.05 was considered significant at 95% confidence level and 0.10 at 90% confidence level. Demographic variables and patient characteristics were summarized descriptively by treatment assignments. Demographic variables include age, gender, weight, and body mass index. Variables measured on a continuous scale, such as the patient’s age at the time of enrolment, the number of non-missing observations (n), mean, and SEM, were tabulated by treatment assignments. All available data were used in the analyses. Each group was summarized individually. Data not available was assessed as “missing values,” and the observed population only were evaluated. The statistical analysis was processed with GraphPad Prism 9.0.2 (GraphPad, San Diego, CA).

## RESULTS

### Patient Enrollment and Demographics

This study was conducted in 14 Emergency Room/Intensive Care Units across India. Patients with hypovolemic shock due to blood loss or fluid loss with SBP ≤ 90 mmHg and lactate level indicative (≥2 mmol/L) of shock at presentation and who continued receiving standard shock treatment were included. A total of 197 patients were assessed, and 105 patients met the eligibility criteria and were enrolled in the study. Out of 105 patients, 71 were randomized in the centhaquine group and 34 in the control group. In the centhaquine group, 1 patient withdrew consent, and 2 patients were excluded by the investigator (one patient was diagnosed with fulminant tuberculosis, and another patient was diagnosed with refractory septic shock). A total of 34 patients (22 male and 12 female) in control and 68 patients (41 male and 27 female) in the centhaquine group completed the study (Figure 1). The average age of patients was 36.50 years in the control group and 42.81 years in the centhaquine group (Table 1). The difference in the means of age (6.309 ± 3.639, two-tailed t-test, 95% CI –0.938 to 13.56, p=0.087) between the two groups indicated that average age was higher in the centhaquine group than the control group (not significant at 95% but significant at 90% confidence interval).

**Figure 1:**
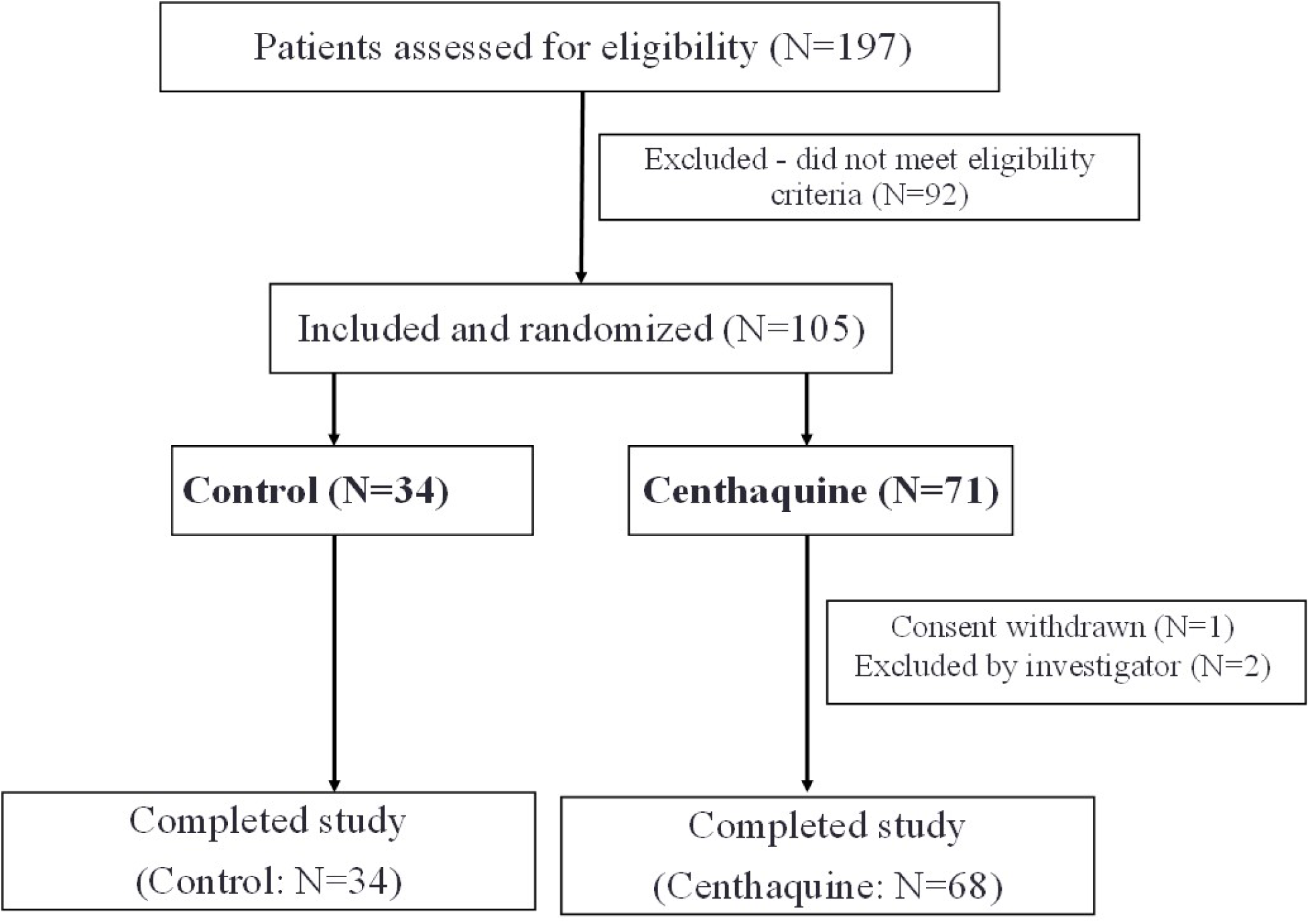
Patient enrolment, randomization, and trial completion.

**Table 1.** Baseline characteristics of patients

|  | **Control (N=34)** | **Centhaquine (N=68)** |
| --- | --- | --- |
| **Age (years)** | 36.50 ± 2.81 | 42.81 ± 2.31 |
| **Body weight (Kg)** | 56.74 ± 1.62 | 58.90 ± 1.37 |
| **Body mass index (Kg/m<sup>2</sup>)** | 21.56 ± 0.59 | 22.72 ± 0.51 |
| **Sex** |  |  |
| *Men* | 22 (64.71 %) | 41 (60.29 %) |
| *Women* | 12 (35.29 %) | 27 (39.71 %) |
| **Medical history** |  |  |
| *Hypertension* | 03 (8.82%) | 01 (1.47%) |
| *Diabetes* | 04 (11.76%) | 03 (4.41%) |
| *Renal disorders* | 01 (2.94%) | 03 (4.41%) |
| *Respiratory disease* | 01 (2.94%) | 02 (2.94%) |
| *Ischemic heart disease* | --- | 01 (1.47%) |
| *Liver fibrosis* | 01 (2.94%) | --- |
| *Hepatitis* | 01 (2.94%) | --- |
| *Pre-eclampsia* | --- | 01 (2.94%) |
| **Reason for hypovolemic shock** |  |  |
| *Trauma* | 10 (29.41 %) | 32 (47.06 %) |
| *Post-surgery* | 05 (14.71 %) | 03 (4.41 %) |
| *Post-partum haemorrhage* | 02 (5.88 %) | 06 (8.82 %) |
| *Vaginal bleeding* | 02 (5.88 %) | 07 (10.29 %) |
| *Gastroenteritis* | 15 (44.12 %) | 20 (29.41 %) |
| **Clinical factors** |  |  |
| *Systolic blood pressure (mmHg)* | 83.62 ± 1.48 | 83.52 ± 1.13 |
| *Diastolic blood pressure (mmHg)* | 53.00 ± 1.47 | 55.82 ± 1.21 |
| *Heart rate (beats/min)* | 115.15 ± 3.32 | 107.76 ± 2.61 |
| *Shock index* | 1.39 ± 0.05 | 1.32 ± 0.05 |
| *Respiratory rate (breaths/min)* | 24.88 ± 1.32 | 24.00 ± 0.86 |
| *Body temperature (°C)* | 36.76 ± 0.07 | 36.80 ± 0.06 |
| *Blood lactate (mmol/L)* | 4.13 ± 0.40 | 4.50 ± 0.29 |
| *Base deficit (mmol/L)* | -7.36 ± 1.07 | -7.58 ± 0.56 |
| *Haemoglobin (g/dL)* | 10.41 ± 0.47 | 9.64 ± 0.35 |
| *Haematocrit (%)* | 31.90 ± 1.56 | 29.48 ± 1.09 |
| *Creatinine (mg/dL)* | 1.28 ± 0.13 | 1.22 ± 0.08 |
| *Glomerular filtration rate (mL/min/1.73 m<sup>2</sup>)* | 78.84 ± 5.42 | 77.41 ± 4.30 |
| *Glasgow Coma Scale* | 14.29 ± 0.31 | 13.40 ± 0.42 |
| *Acute respiratory distress syndrome* | 0.15 ± 0.05 | 0.18 ± 0.04 |
| *pH* | 7.32 ± 0.02 | 7.34 ± 0.01 |
| *pCO<sub>2</sub> (mmHg)* | 33.65 ± 1.36 | 31.98 ± 0.90 |
| *paO<sub>2</sub> (mmHg)* | 117.04 ± 9.39 | 113.48 ± 5.62 |

Data are presented as mean ± standard error of the mean.

### Patient Assessment at the Time of Inclusion

History of hypertension, diabetes, renal and hepatic disorders, and other medical conditions were similar in both groups (Table 1). However, trauma was the predominant cause of hypovolemic shock in the centhaquine group (47.06%) than in the control group (29.41%) (Chi-square test, OR 2.133, 95% CI 0.9146 to 5.269, p=0.0878). On the other hand, gastroenteritis was the leading cause of hypovolemic shock in the control group (44.12%) than the centhaquine group (29.41%)(Chi-square test, OR 0.528, 95% CI 0.222 to 1.264, p=0.1403). Other reasons for hypovolemic shock such as post-surgical blood loss, post-partum hemorrhage, vaginal bleeding did not differ in both groups. Baseline clinical parameters of blood pressure, heart rate, shock index, respiratory rate, base deficit, and body temperature were similar in centhaquine and control groups (Table 1). Blood lactate level was 4.13 ± 0.40 mmol/L in the control group and 4.50 ± 0.29 mmol/L in the centhaquine group. Hemoglobin level was slightly lower in the centhaquine group 9.64 ± 0.35 g/dL than the control group 10.41 ± 0.47 g/dL; similarly, hematocrit was a little lower in the centhaquine group 29.48 ± 1.09 % than the control group 31.90 ± 1.56 % (Table 1). The difference between the means of hemoglobin (−0.769 ± 0.590, two-tailed t-test, 95% CI -1.946 to 0.408, p=0.1968) and hematocrit levels (−2.419 ± 1.899, two-tailed t-test, 95% CI -6.213 to 1.374, p=0.2073) between control and centhaquine groups suggest that the blood loss is slightly more in the centhaquine group than the control group. Creatinine levels and glomerular filtration rate were similar in the two groups. No difference in the Glasgow coma scale and ARDS was observed between the two groups. Details of the cause of hypovolemic shock and any surgical procedure performed during hospitalization of the individual patient are in Supplementary Table 2 for the control group and Supplementary Table 3 for the centhaquine group.

### Total fluids, blood and blood products, and vasopressors

The total volume of fluids (crystalloids, colloids) administered before randomization was similar in the control and centhaquine groups (Table 2). The total blood and blood product administered before randomization were 0.051 ± 0.04 L in the control and 0.12 ± 0.04 L in the centhaquine group. The amount of vasopressors administered before randomization were similar in both groups (Table 2). After randomization, the number of doses of study drug administered in the control group averaged 1.47 ± 0.19 per patient, and in the centhaquine group, it was 1.27 ± 0.03 per patient (difference between the means -0.187 ± 0.204, two-tailed t-test, 95% CI - 0.599 to 0.225, p=0.3657) during 48 hours of resuscitation. Following randomization, the total amount of fluids administered in 48 hours was 4.61 ± 0.30 L in the centhaquine group and 4.65 ± 0.37 L in the control group (Figure 2 and Table 2). The amount of blood and blood products infused during the first 24 and 48 hours were similar in both groups (Table 2 and Figure 2). A total dose of vasopressors administered in 48 hours in the control group (4.40 ± 2.41 mg) appeared to be higher compared to the centhaquine group (2.76 ± 1.28 mg); however, this difference (difference between means of control and centhaquine -1.645 ± 2.728, 95% CI - 7.119 to 3.829, p=0.549) was not statistically significant (Figure 2 and Table 2). During the first 48 hours of resuscitation, urine output was similar in both groups (Figure 2). Pharmacological treatment provided to patients in control and centhaquine groups is shown in supplementary table 4. Both groups received comparatively similar pharmacological agents (Supplementary Table 4).

**Figure 2:**
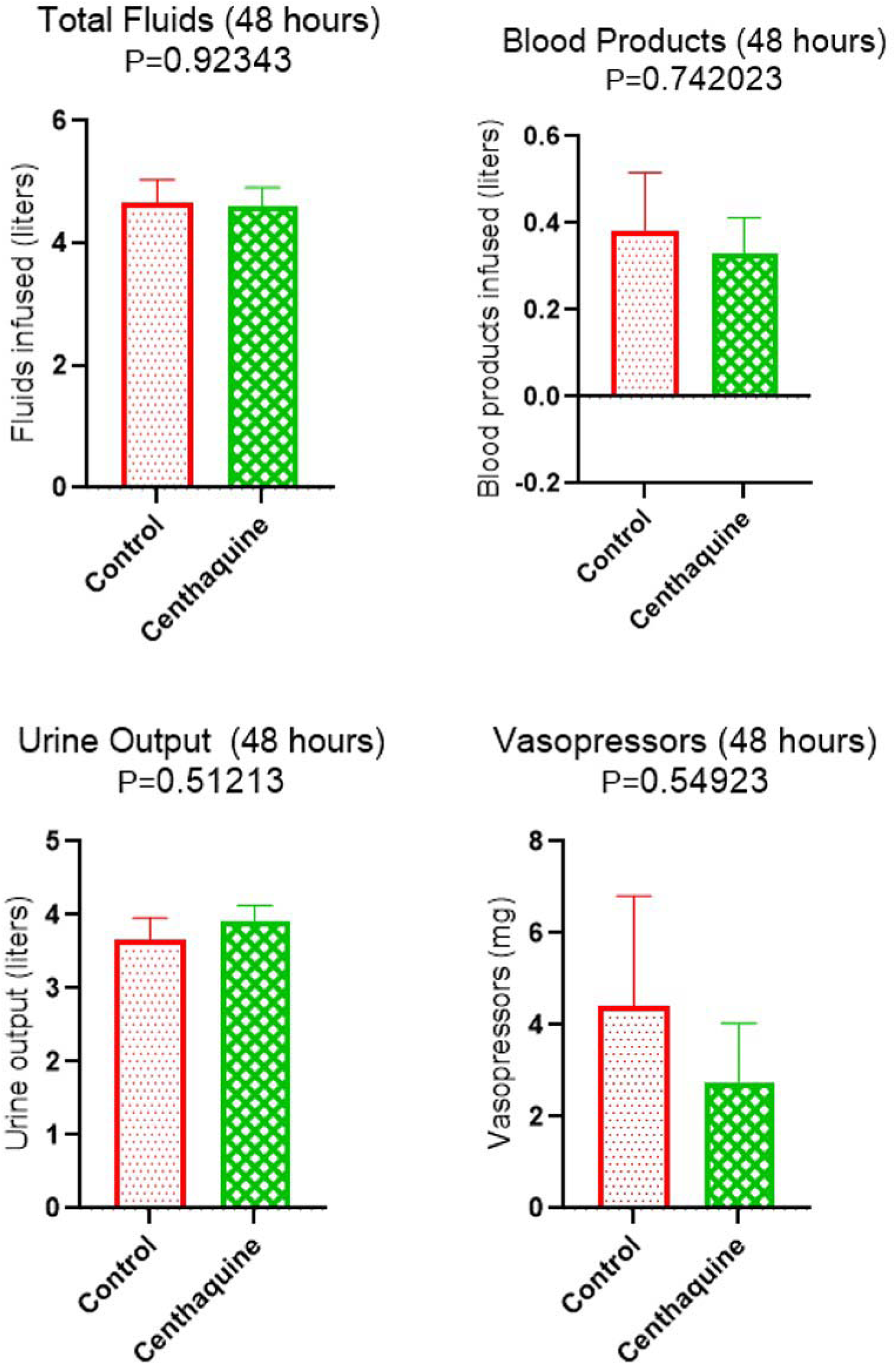
Total volume of fluid, blood products, and vasopressors administered during the first 48 hours in the control and centhaquine group of patients. Total urine output in the first 48 hours in the control and centhaquine group of patients. Data presented as the mean ± standard error.

**Table 2.** Standard treatment administered to the patients before randomization and during the first 24 hours and 48 hours of resuscitation

| **Pre-dose (Before Randomization)** |  |  |  |  |  |  |  |  |  |  |
| --- | --- | --- | --- | --- | --- | --- | --- | --- | --- | --- |
| Group | Fluids (L) |  |  | Blood Products (L) |  |  |  |  |  | Vasopressors (mg) |
|  | Crystalloids | Colloids | **Total Volume** | Blood | Packed red blood cells | Fresh frozen plasma | Platelets | Cryoprecipitate | **Total Volume** | **Total Dose** |
| Control | 0.76±0.15 | 0.044±0.02 | **0.81±0.17** | --- | 0.033±0.02 | 0.021±0.021 | --- | --- | **0.051±0.04** | **0.23±.12** |
| Centaquine | 0.75±0.09 | 0.057±0.02 | **0.80±0.11** | 0.031±0.021 | 0.072±0.02 | 0.008±0.008 | --- | 0.011±0.011 | **0.120±0.04** | **0.37±0.17** |
| **Post-dose (First 24 hours of Resuscitation)** |  |  |  |  |  |  |  |  |  |  |
| Control | 2.86±0.27 | 0.079±0.04 | **2.94±0.31** | 0.061±0.033 | 0.113±0.04 | 0.079±0.056 | --- | --- | **0.254±0.09** | **3.83±2.31** |
| Centaquine | 2.77±0.20 | 0.032±0.02 | **2.79±0.23** | 0.005±0.005 | 0.181±0.04 | 0.054±0.027 | 0.0093±0.006 | --- | **0.249±0.07** | **2.26±1.20** |
| **Post-dose (First 48 hours of Resuscitation)** |  |  |  |  |  |  |  |  |  |  |
| Control | 4.58±0.37 | 0.079±0.04 | **4.65±0.37** | 0.074±0.043 | 0.162±0.05 | 0.136±0.078 | 0.010±0.010 | --- | **0.382±0.14** | **4.40±2.41** |
| Centaquine | 4.57±0.29 | 0.038±0.02 | **4.61±0.30** | 0.005±0.005 | 0.259±0.05 | 0.054±0.027 | 0.0093±0.006 | --- | **0.328±0.08** | **2.76±1.28** |

The fluids administered to the patients were crystalloids (normal saline, dextrose, sodium bicarbonate, PlasmaLyte and lactated ringer) and colloids (albumin, Haemaccel and Gelofusine). The vasopressors used were adrenaline, noradrenaline, and vasopressin.

### Time in hospital, ICU, and on the ventilator

Duration of hospital stay in control was 6.93 ± 1.37 days, while patients in the centhaquine group stayed in the hospital for 8.66 ± 1.17 days. Patients in the centhaquine groups stayed in the hospital for 1.729 ± 1.804 (two-tailed t-test, 95% CI -1.874 to 5.332, p=0.3414) days longer than the patients in the control group. Duration of stay in the intensive care unit in the control group was 2.19 ± 0.43 days, while patients in the centhaquine group stayed in the intensive care unit for 2.89 ± 0.67 days. Patients in the centhaquine group were 0.7008 ± 0.8002 (two-tailed t-test, 95% CI -0.8898 to 2.291, p=0.3836) days longer in the intensive care unit than the control group of patients. However, percent time patients spent in the intensive care unit during their stay in the hospital was similar in both the groups 46.61 ± 7.95 in the control group and 46.03 ± 5.72 in the centhaquine group (difference between means -0.5064 ± 9.801, 95% CI -20.14 to 19.13, p=0.9590). Time on the ventilator was 0.08 ± 0.05 days in the control group and 0.86 ± 0.5 days in the centhaquine group.

### Systemic Hemodynamics

Centhaquine increased SBP more significantly in the initial hours of resuscitation than control (Figure 3). At 15 minutes of resuscitation mean difference in SBP from baseline was 3.618 mmHg (95% CI -3.109 to 10.34, p=0.9464) in the control group while it was 5.652 mmHg (95% CI 0.3844 to 10.92, p=0.0226) in the centhaquine group. The mean difference in SBP from baseline at 30 min of resuscitation was 6.324 mmHg (95% CI 0.4033 to 13.05, p=0.0915) in the control group, while it was 8.621 mmHg (95% CI 3.354 to 13.89, p<0.0001) in the centhaquine group. An increase in SBP from baseline was more with centhaquine than control at 1, 12, 24, and 48 hours of resuscitation. The mean difference in SBP from baseline at 1 hour of resuscitation was 11.00 mmHg (95% CI 4.273 to 17.73) in the control group while 15.21 mmHg (95% CI 9.945 to 20.48) in the centhaquine group. At 12 hours, it was 23.99 mmHg (95% CI 17.21 to 30.77) in control and 26.38 mmHg (95% CI 21.09 to 31.67) in centhaquine groups, and at 24 hours, it was 27.60 mmHg (95% CI 20.82 to 34.38) in control and 32.03 mmHg (95% CI 26.72 to 37.35) in the centhaquine group. The SBP at 48 hours after resuscitation was 116.65 ± 1.63 mmHg in control and 119.81 ± 1.78 mmHg in the centhaquine group (Figure 3 and Table 3). The number of patients with SBP of >90 mmHg at 12 hours of resuscitation was higher in the centhaquine (96.92%) group than the control (87.50%) group (Chi-square test, OR 4.50, 95% CI 0.983 to 24.31, p=0.0701). At 24 hours of resuscitation, patients with SBP of ≥110 mmHg were significantly higher in the centhaquine (79.69%) group than the control (60.61%) group (Chi-square test, OR 2.55, 95% CI 1.033 to 6.391, p=0.0444). SBP of ≥120 mmHg was in 46.88% and 56.25% patients of control and centhaquine group, respectively (Chi-square test, OR 1.457, 95% CI 0.622 to 3.506, p=0.3855) (Figure 3).

**Figure 3:**
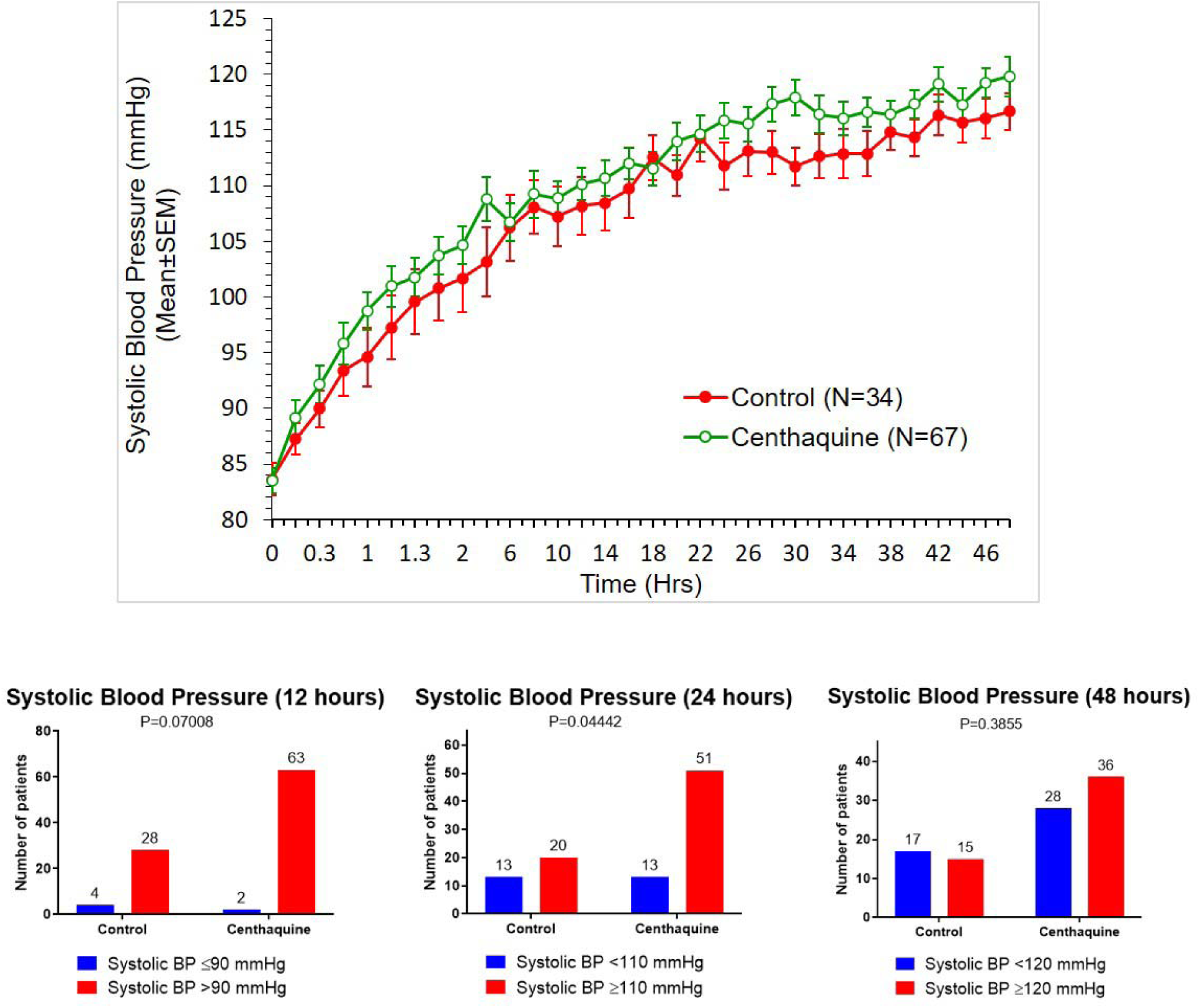
Systolic blood pressure during the first 48 hours in the control and centhaquine group of patients. The upper panel shows data as the mean ± standard error. The lower panel indicates the number of patients with improved systolic blood pressure at 12, 24, and 48 hours of resuscitation.

**Table 3.**
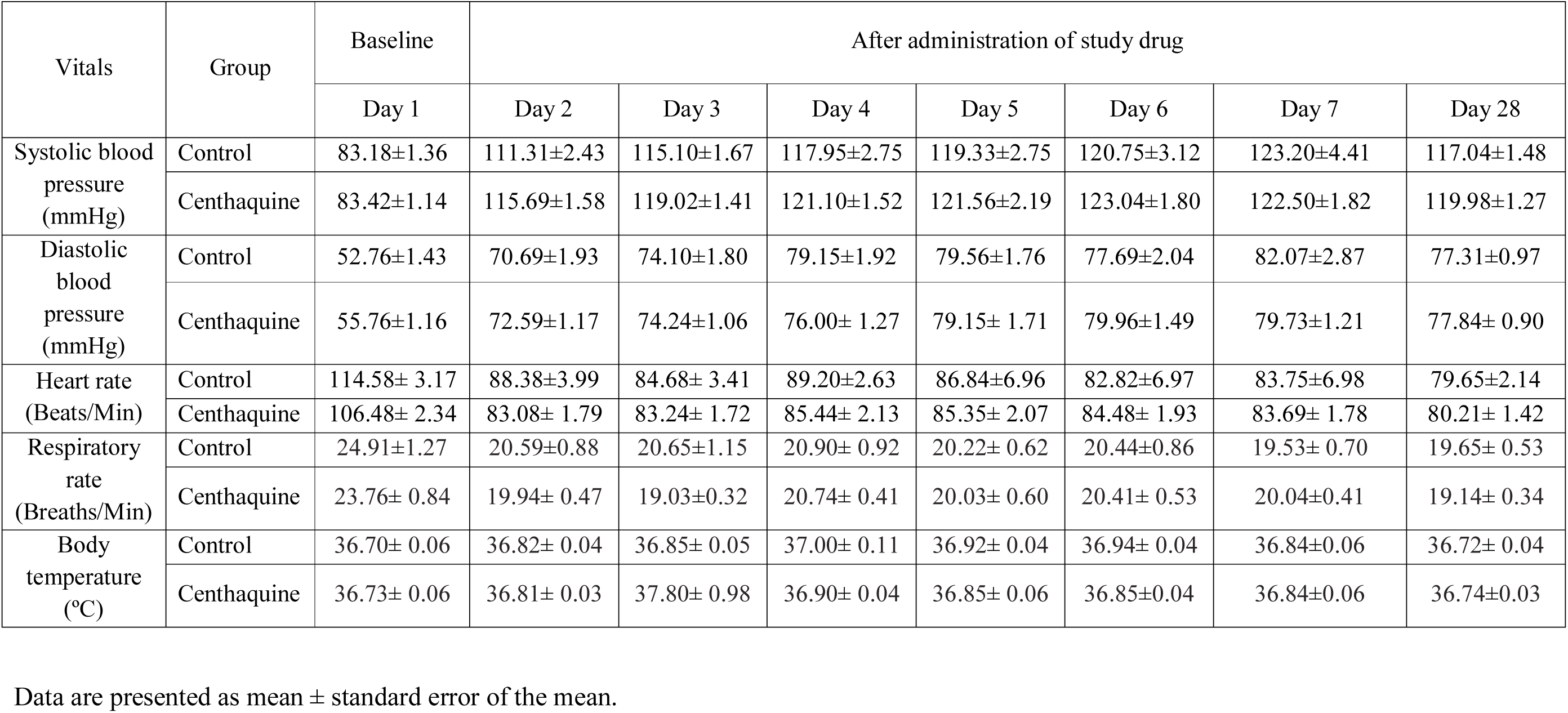
Patients vitals recorded from day 1 (baseline) through day 28

| Vitals | Group | Baseline | After administration of study drug |  |  |  |  |  |  |
| --- | --- | --- | --- | --- | --- | --- | --- | --- | --- |
|  |  | Day 1 | Day 2 | Day 3 | Day 4 | Day 5 | Day 6 | Day 7 | Day 28 |
| Systolic blood pressure (mmHg) | Control | 83.18±1.36 | 111.31±2.43 | 115.10±1.67 | 117.95±2.75 | 119.33±2.75 | 120.75±3.12 | 123.20±4.41 | 117.04±1.48 |
|  | Cenchaquine | 83.42±1.14 | 115.69±1.58 | 119.02±1.41 | 121.10±1.52 | 121.56±2.19 | 123.04±1.80 | 122.50±1.82 | 119.98±1.27 |
| Diastolic blood pressure (mmHg) | Control | 52.76±1.43 | 70.69±1.93 | 74.10±1.80 | 79.15±1.92 | 79.56±1.76 | 77.69±2.04 | 82.07±2.87 | 77.31±0.97 |
|  | Cenchaquine | 55.76±1.16 | 72.59±1.17 | 74.24±1.06 | 76.00± 1.27 | 79.15± 1.71 | 79.96±1.49 | 79.73±1.21 | 77.84± 0.90 |
| Heart rate (Beats/Min) | Control | 114.58± 3.17 | 88.38±3.99 | 84.68± 3.41 | 89.20±2.63 | 86.84±6.96 | 82.82±6.97 | 83.75±6.98 | 79.65±2.14 |
|  | Cenchaquine | 106.48± 2.34 | 83.08± 1.79 | 83.24± 1.72 | 85.44± 2.13 | 85.35± 2.07 | 84.48± 1.93 | 83.69± 1.78 | 80.21± 1.42 |
| Respiratory rate (Breaths/Min) | Control | 24.91±1.27 | 20.59±0.88 | 20.65±1.15 | 20.90± 0.92 | 20.22± 0.62 | 20.44±0.86 | 19.53± 0.70 | 19.65± 0.53 |
|  | Cenchaquine | 23.76± 0.84 | 19.94± 0.47 | 19.03±0.32 | 20.74± 0.41 | 20.03± 0.60 | 20.41± 0.53 | 20.04±0.41 | 19.14± 0.34 |
| Body temperature (°C) | Control | 36.70± 0.06 | 36.82± 0.04 | 36.85± 0.05 | 37.00± 0.11 | 36.92± 0.04 | 36.94± 0.04 | 36.84±0.06 | 36.72± 0.04 |
|  | Cenchaquine | 36.73± 0.06 | 36.81± 0.03 | 37.80± 0.98 | 36.90± 0.04 | 36.85± 0.06 | 36.85±0.04 | 36.84±0.06 | 36.74±0.03 |

Data are presented as mean ± standard error of the mean.

An increase in DBP was similar in both the control and centhaquine groups (Figure 4). The mean difference in DBP at 15 minutes from the baseline was 4.568 mmHg (95% CI -0.613 to 9.75, p=0.1575) in the control group, while it was 3.877 mmHg (95% CI -0.4437 to 8.198, p=0.1371) in the centhaquine group. At 30 minutes of resuscitation mean difference in DBP from baseline was 5.629 mmHg (95% CI 0.4472 to 10.81, p=0.0195) in the control group while it was 5.738 mmHg (95% CI 1.418 to 10.06, p=0.0009) in the centhaquine group. An increase in DBP from baseline was similar in centhaquine than control groups at 1, 12, 24, and 48 hours of resuscitation. The mean difference in DBP from baseline at 1 hour of resuscitation was 8.794 mmHg (95% CI 3.653 to 13.94) in the control group, while it was 7.769 mmHg (95% CI 3.449 to 12.09) in the centhaquine group, and at 12 hours, it was 15.56 mmHg (95% CI 10.38 to 20.74) in control and 15.12 mmHg (95% CI 10.79 to 19.44) in centhaquine groups. At 24 hours, the difference was 16.71 mmHg (95% CI 11.53 to 21.89) in control and 16.50 mmHg (95% CI 12.16 to 20.85) in the centhaquine group. The SBP 48 hours after resuscitation was 74.594 ± 1.272 mmHg in control and 76.469 ± 0.938 mmHg in the centhaquine group (Figure 4). The number of patients with DBP of >65 mmHg at 12 hours of resuscitation was 72.31% in the centhaquine group and 60.61% in the control (60.61%) group (Chi-square test, OR 1.697, 95% CI 0.7333 to 4.259, p=0.2391). At 24 hours of resuscitation, patients with DBP of ≥70 mmHg were significantly higher in the centhaquine (76.56%) group than the control (51.52%) group (Chi-square test, OR 3.07, 95% CI 1.215 to 7.266, p=0.0122). A DBP of ≥80 mmHg was in 31.25% and 50.00% patients of the control and centhaquine group, respectively (Chi-square test, OR 2.200, 95% CI 0.9107 to 5.588, p=0.0809) (Figure 4).

**Figure 4:**
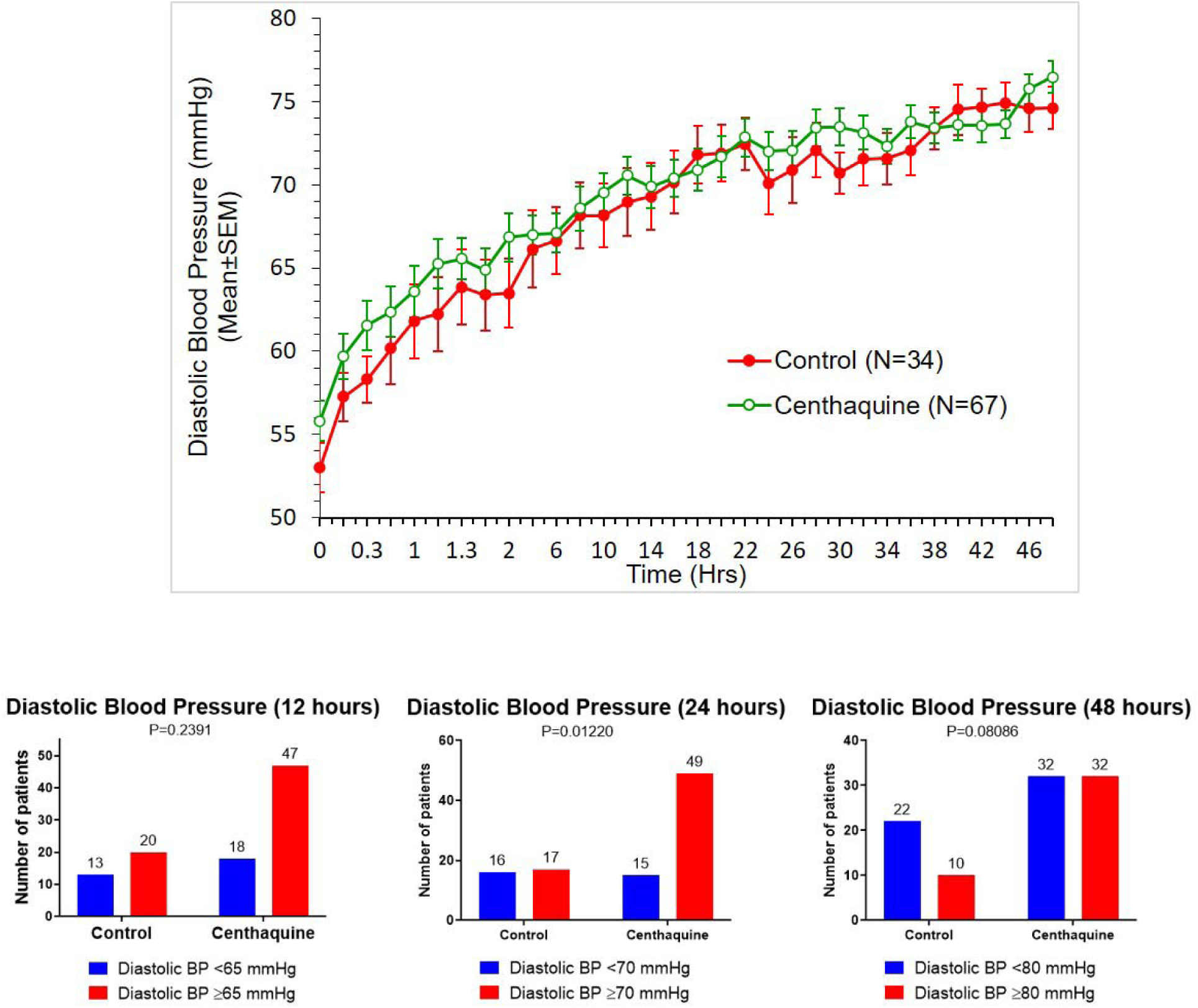
Diastolic blood pressure during the first 48 hours in the control and centhaquine group of patients. The upper panel shows data as the mean ± standard error. The lower panel indicates the number of patients with improved diastolic blood pressure at 12, 24, and 48 hours of resuscitation.

Centhaquine increased pulse pressure more significantly in the initial hours of resuscitation than control. At 15 minutes of resuscitation mean difference in pulse pressure from baseline was -0.7576 mmHg (95% CI -6.094 to 4.579, p>0.9999) in the control group while it was 1.862 mmHg (95% CI -2.541 to 6.264, p=0.9981) in the centhaquine group. The difference at 30 min of resuscitation was 0.7878 mmHg (95% CI -4.549 to 6.124, P>0.999) in the control group, while it was 2.862 mmHg (95% CI -1.541 to 7.264, p=0.7241) in the centhaquine group. At 45 minutes of resuscitation, it was 2.618 mmHg (95% CI -2.678 to 7.913, p=0.9804) in the control group and 5.785 mmHg (95% CI 1.382 to 10.19, p=0.0011) in the centhaquine group. At 60 minutes, the difference was 2.206 mmHg (95% CI -3.089 to 7.501, p=0.9985) in control and 7.523 mmHg (95% CI 3.121 to 11.93, P<0.0001) in centhaquine groups. Then at 90 minutes, the difference was 5.509 mmHg (95% CI - 0.2365 to 10.35, p=0.0788) in control and 8.646 mmHg (95% CI 4.244 to 13.05, P<0.0001) in centhaquine groups. An increase in pulse pressure from a baseline of 30.61 ± 1.228 to 42.06 ± 1.13 mmHg (11.03 mmHg increase, 95% CI 5.650 to 16.41) at 48 hours of resuscitation was observed in the control group, while in the centhaquine group, it increased from a baseline of 28.37 ± 1.05 to 43.34 ± 1.44 mmHg (14.68 mmHg increase, 95% CI 10.25 to 19.10) at 48 hours of resuscitation.

The area under the curve (AUC) of the mean difference in SBP, DBP, and pulse pressure from baseline to various time intervals from baseline to 48 hours was determined (Figure 5). AUC for SBP was 709.6 in the control group and 801.8 in the centhaquine group, indicating a 12.99% increase in AUC than the control group. In contrast, AUC for DBP was 464.9 in the control group and 430.3 in the centhaquine group, indicating a decrease in AUC of 7.44% in the centhaquine than the control group. AUC of pulse pressure was 245.1 in the control group and 363.1 in the centhaquine group, indicating an increase in AUC of 48.14% in the centhaquine than the control group (Figure 5).

**Figure 5:**
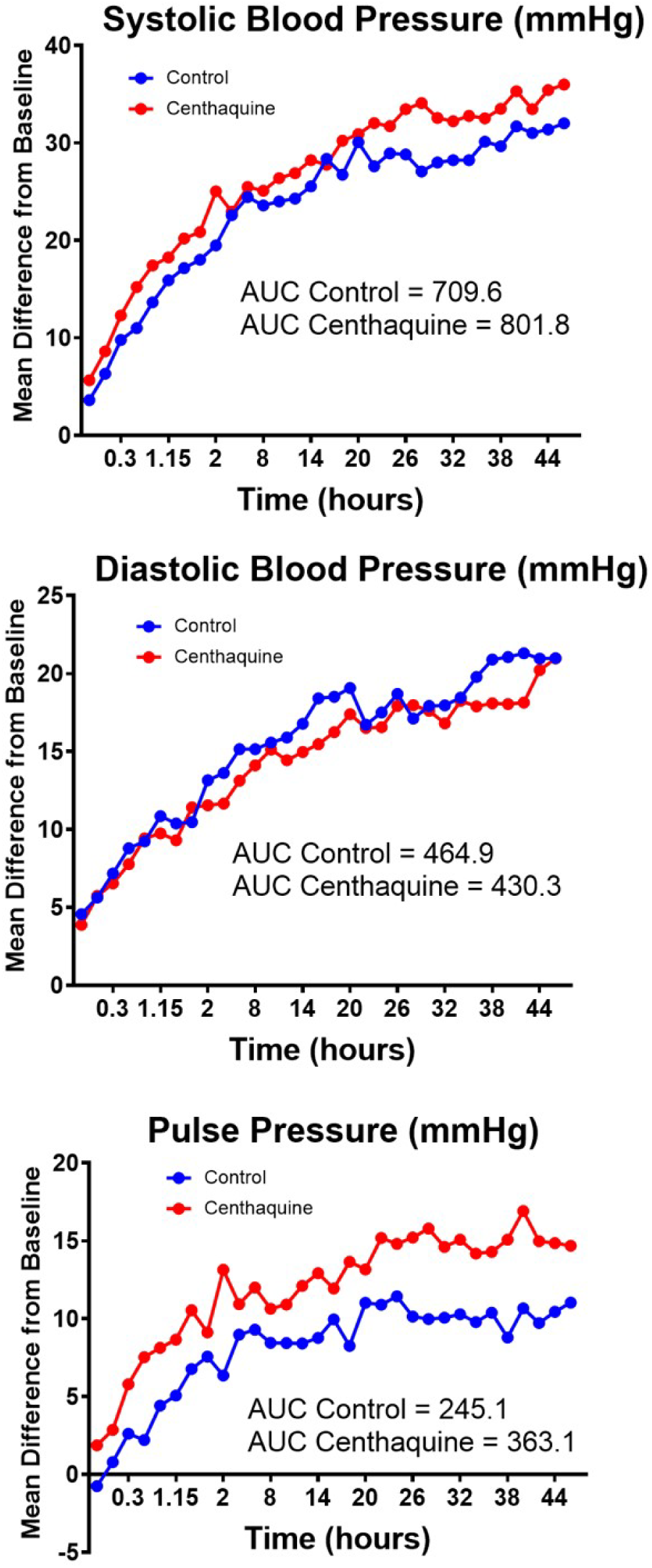
Mean difference from baseline at various time intervals plotted to determine the area under the curve (AUC) for systolic blood pressure, diastolic blood pressure, and pulse pressure. AUC for systolic blood pressure was higher by 12.99%, diastolic blood pressure lower by 7.44%, and pulse pressure higher by 48.14% in the centhaquine group than the control group. A significant increase in pulse pressure in the centhaquine group highly suggests an increase in stroke volume.

A decrease in heart rate was observed in control and centhaquine patients. Heart rate decreased from a baseline of 114.58 ± 3.17 to 88.38 ± 3.99 beats/min on day 2 of resuscitation in the control group and from a baseline of 106.48 ± 2.34 to 83.08 ± 1.79 beats/min in the centhaquine group (Table 3). SBP, DBP, heart rate, respiratory rate, and body temperature records till 28 days are provided in Table 3.

### Shock index

The mean shock index (SI) at the time of inclusion (0 hours) was 1.390 and 1.320 in control and centhaquine groups, respectively (difference between means 0.070 ± 0.066; 95% CI -0.2022 to 0.06165, p=0.2926), indicating that degree of shock was similar in both groups and was moderate to severe. At one hour of resuscitation, SI decreased to a mean of 1.171 and 1.020 in control and centhaquine groups, respectively, which was significantly lower in the centhaquine group than control (difference between means 0.1505 ± 0.0682; 95% CI -0.2876 to -0.0134, p=0.0321). SI was significantly lower in the centhaquine group at four hours of resuscitation than in control (difference between means 0.1370 ± 0.0677; 95% CI -0.2736 to -0.0003, p=0.0494). SI significantly improved in the centhaquine group in the first 4 hours of resuscitation (Figure 6).

**Figure 6:**
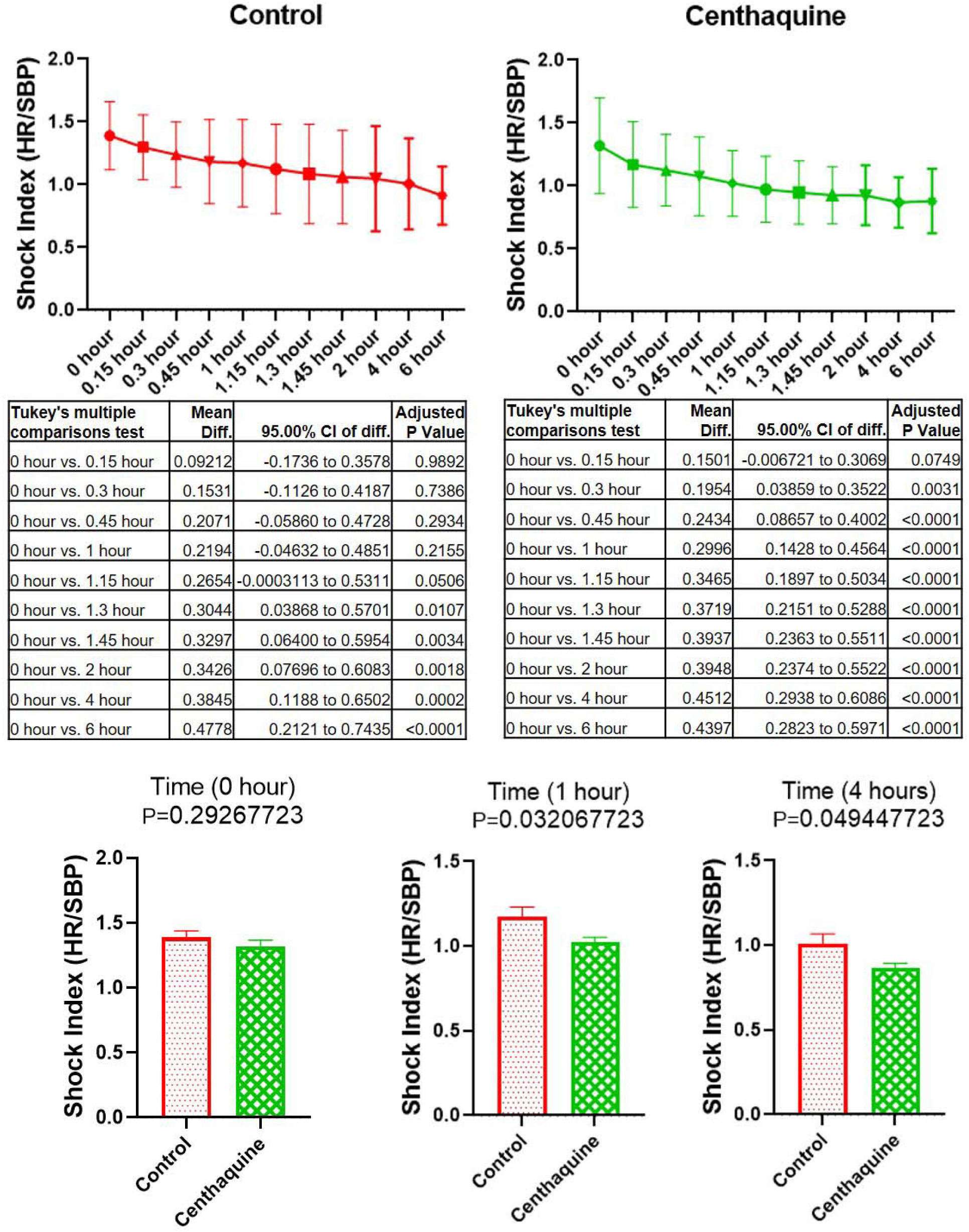
Shock index (heart rate divided by systolic blood pressure), an important indicator of cardiac performance (left ventricular stroke work) in early hemorrhage, was significantly improved by centhaquine in the first 4 hours of resuscitation.

### Blood Lactate Levels

The blood lactate levels in hypovolemic shock patients were high on day 1, ranging from 2.04 to 11.00 mmol/L (mean ± SEM, 4.44 ± 0.29) in the centhaquine group and from 2.04 to 14.12 mmol/L (mean ± SEM, 4.14 ± 0.42) in the control group. Treatment with centhaquine decreased blood lactate levels, as evidenced by blood lactate levels on day 3 that ranged from 0.6 to 4.82 mmol/L (mean ± SEM, 1.43 ± 0.09). Except for one (out of 68 patients), every patient treated with centhaquine had lower blood lactate levels on day 3 compared to day 1. In that patient, blood lactate levels were 2.69 and 4.82 mmol/L on day 1 and day 3, respectively. In the centhaquine group, this patient was the only outlier with no decrease in blood lactate levels. In the control group, blood lactate levels on day 3 ranged from 0.32 to 7.52 mmol/L (mean ± SEM, 1.91 ± 0.26). In this group, two (out of 34) patients had higher blood lactate levels on day 3 than day 1. One patient had blood lactate levels of 4.80 and 5.30 mmol/L on day 1 and day 3, respectively, and the other patient had blood lactate levels of 2.12 and 2.48 mmol/L on day 1 and day 3, respectively. On day 3 of resuscitation, blood lactate levels were lower in the centhaquine group than the control group (difference in the means -0.4828 ± 0.279; 95% CI -1.048 to 0.08253, p=0.0919). The number of patients with blood lactate levels of 1.5 mmol or less was 46.88% in the control group compared to 69.35% in the centhaquine group (OR 2.565, 95% CI 1.047 to 5.873, p=0.0336) (Figure 7).

**Figure 7:**
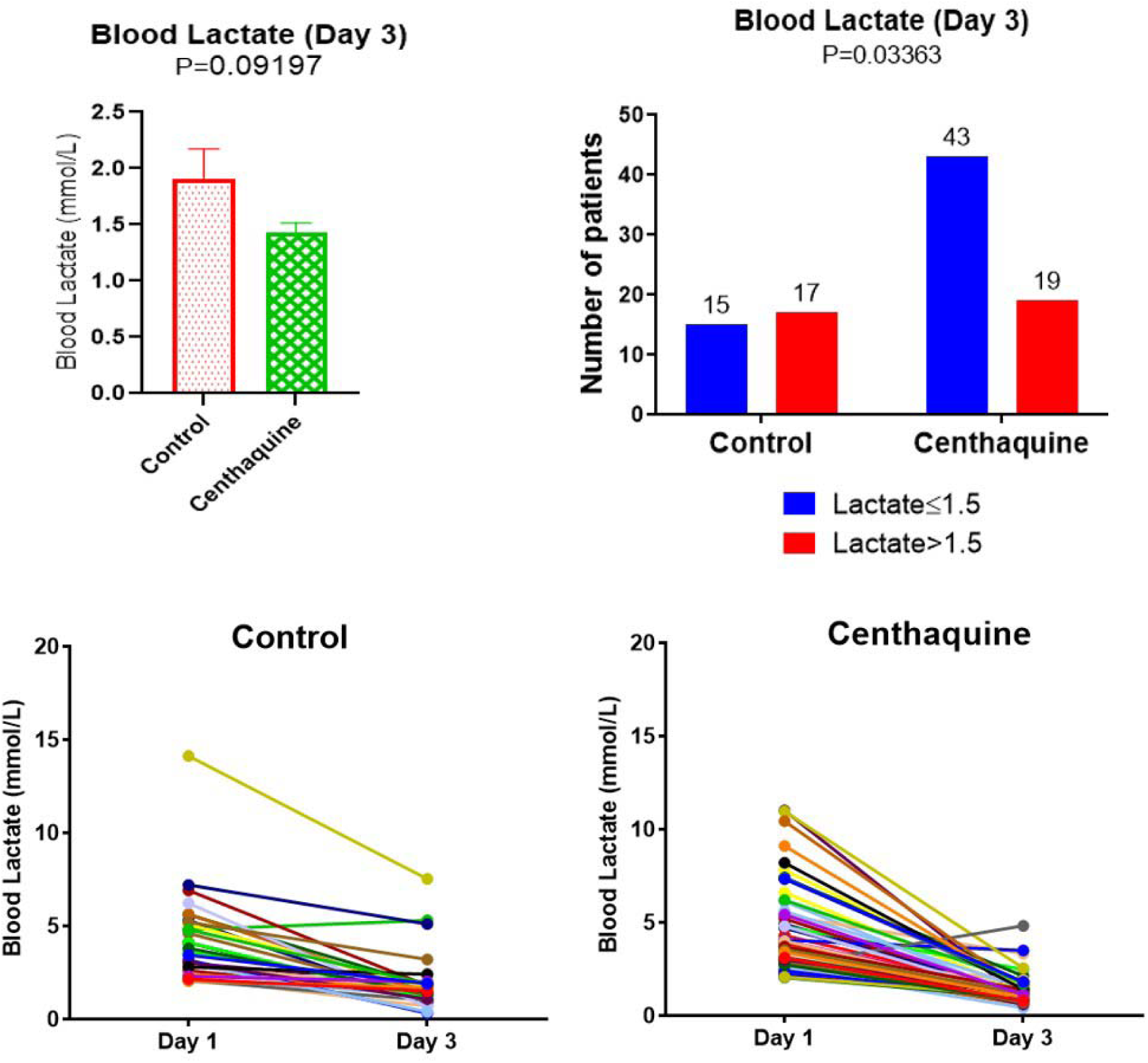
Blood lactate levels in the control and centhaquine group on day 3 of resuscitation are shown in the upper panel. Changes in blood lactate levels following resuscitation of patients with hypovolemic shock in control and centhaquine groups of individual patients are shown in the lower panel.

### Base Deficit

The base deficit ranged from 0.10 to -29.4 in the control group on day 1, and the mean ± SEM was -7.36 ± 0.1.07. The centhaquine group base deficit ranged from - 1.60 to -21.8 mmol/L on day 1 (mean ± SEM -7.58 ± 0.56). An improvement in the base deficit was observed on day 3 of resuscitation and only 4 out of 68 patients (5.88%) treated with centhaquine had a lower base deficit on day 3 compared to day 1, while in the control group, 7 out of 34 patients (20.59%) had a lower base deficit on day 3 compared to day 1. The mean ± SEM of base deficit was -1.84 ± 0.50 on day 3 in the centhaquine group (difference between means from day1 -5.512 ± 0.616; 95% CI -6.744 to -4.280). In the control group, the mean ± SEM of base deficit was -3.33 ± 0.92 on day 3 (difference between means from day 1 -3.520 ± 0.931; 95% CI -5.419 to -1.622). Base-deficit on day 3 of resuscitation improved in patients treated with centhaquine by 1.495 ± 1.045 mmol/L than the control group (95% CI -0.6040 to 3.595, p=0.1587). On day 3 of resuscitation, the number of patients with a base deficit of less than minus 2 was 43.75% in the control group compared to 69.84% in the centhaquine group (OR 2.977, 95% CI 1.223 to 6.907, p=0.0137) (Figure 8).

**Figure 8:**
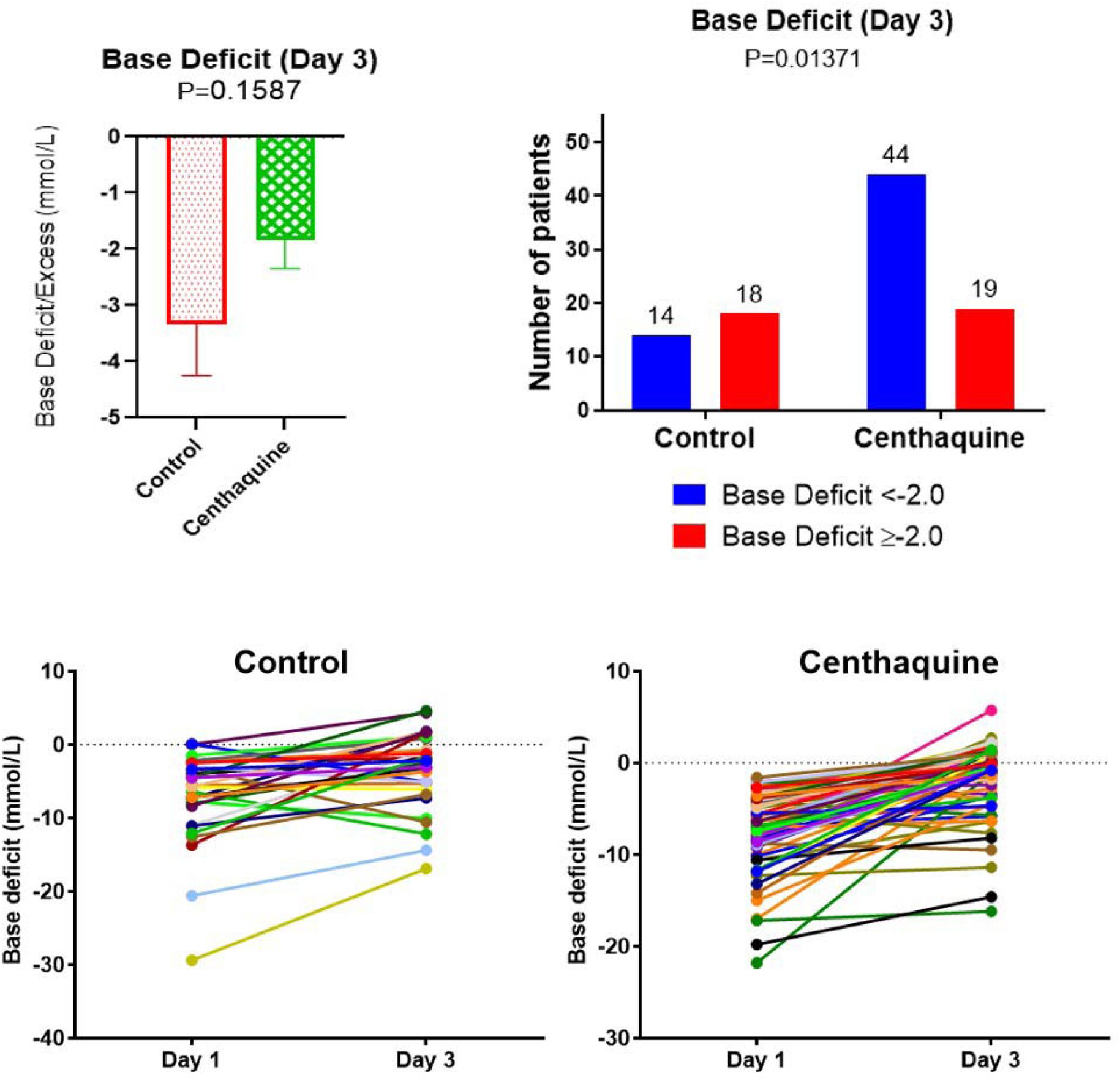
Base deficit in control and centhaquine group on day 3 of resuscitation is shown in the upper panel. Changes in base deficit following resuscitation of patients with hypovolemic shock in control and centhaquine groups of individual patients are shown in the lower panel.

### ARDS and MODS

ARDS was compared between day 1 (before resuscitation) and day 3 of resuscitation. In patients receiving the standard treatment in the control group, the difference between means from day 1 to day 3 was -0.06247 ± 0.05030 (95% CI - 0.1622 to -0.0373, p=0.2171). Conversely, in the centhaquine treated group of patients, the ARDS difference between means from day 1 to day 3 was -0.09927 ± 0.04891 (95% CI -0.1963 to -0.00228, p=0. 0449). These results indicate that centhaquine treatment significantly improved ARDS following resuscitation, whereas there was a minor improvement in the control group (Figure 9 and Table 4).

**Figure 9:**
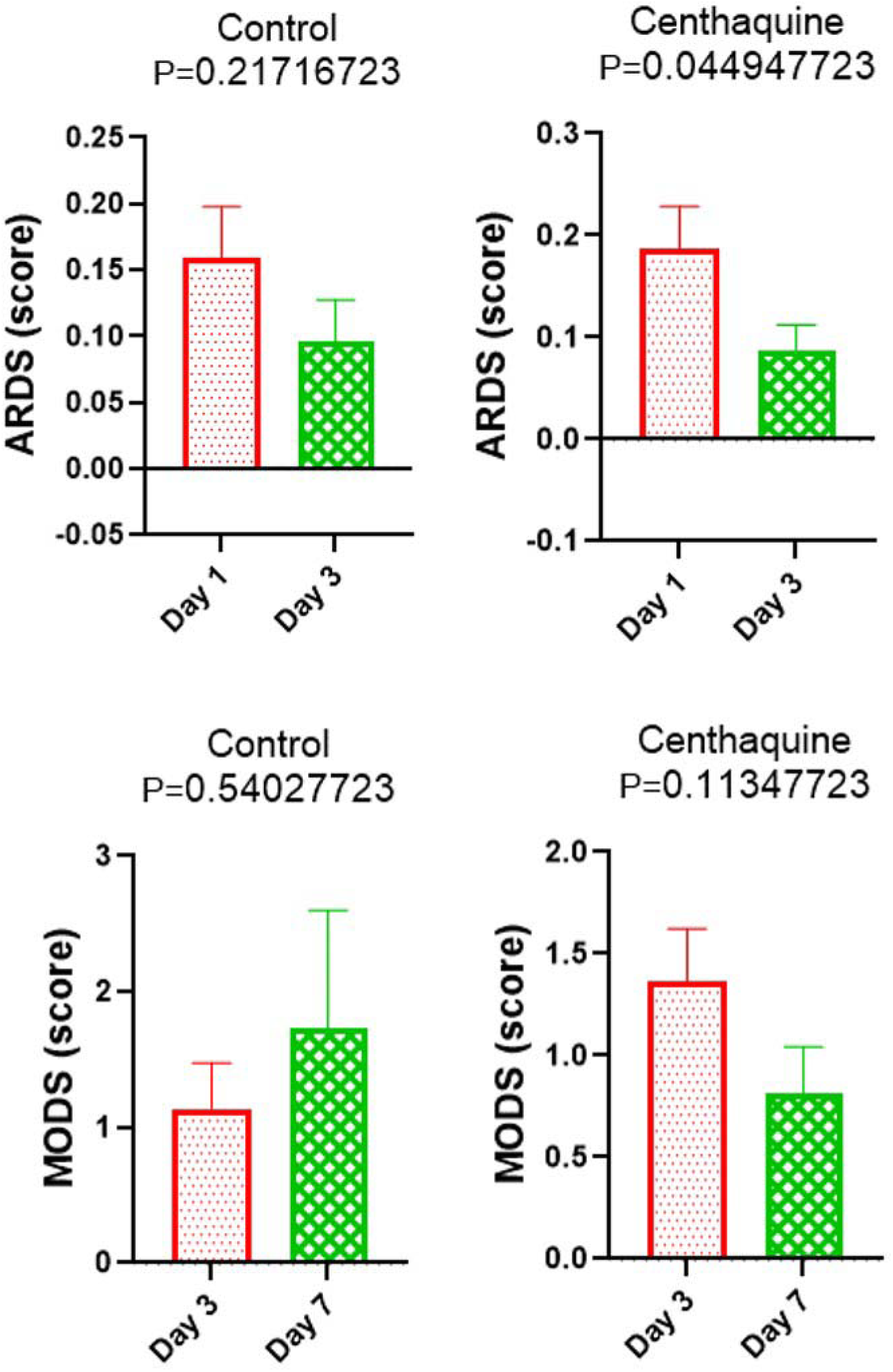
Acute Respiratory Distress Syndrome (ARDS) was compared between day 1 (before resuscitation) and day 3 of resuscitation. Centhaquine treatment significantly improved ARDS following resuscitation, whereas there was a minor improvement in the control group. Multiple Organ Dysfunction Score (MODS) was compared between day 3 and day 7 of resuscitation. MODS in the control group worsened from 1.138 to 1.727, while in the centhaquine group, it improved from 1.367 to 0.8182.

**Table 4.** Patients GCS, MODS, and ARDS, recorded through day 28

| Score | Group | Day 1 | Day 2 | Day 3 | Day 4 | Day 5 | Day 6 | Day 7 | Day 28 |
| --- | --- | --- | --- | --- | --- | --- | --- | --- | --- |
| GCS | Control | 14.29 ± 0.31 | 14.66 ± 0.20 | 14.97 ± 0.03 | 15.00 ± 0.00 | 15.00 ± 0.00 | 15.00 ± 0.00 | 15.00 ± 0.00 | 15.00 ± 0.00 |
|  | Cenchaquine | 13.40 ± 0.42 | 14.34 ± 0.22 | 14.21 ± 0.29 | 14.23 ± 0.35 | 14.29 ± 0.39 | 14.26 ± 0.46 | 14.23 ± 0.46 | 14.91 ± 0.09 |
| MODS | Control |  |  | 1.14 ± 0.34 | 1.65 ± 0.50 | 1.20 ± 0.34 | 1.54 ± 0.69 | 1.73 ± 0.87 | 0.25 ± 0.12 |
|  | Cenchaquine |  |  | 1.37 ± 0.26 | 1.82 ± 0.41 | 1.34 ± 0.26 | 1.09 ± 0.29 | 0.82 ± 0.22 | 0.33 ± 0.10 |
| ARDS | Control | 0.15 ± 0.05 | 0.02 ± 0.01 | 0.08 ± 0.04 | 0.05 ± 0.04 | 0.01 ± 0.01 | 0.06 ± 0.05 | 0.06 ± 0.06 | 0.03 ± 0.03 |
|  | Cenchaquine | 0.18 ± 0.04 | 0.13 ± 0.03 | 0.08 ± 0.02 | 0.13 ± 0.04 | 0.05 ± 0.02 | 0.03 ± 0.02 | 0.08 ± 0.04 | 0.03 ± 0.02 |

Data are presented as mean ± standard error of the mean.

MODS was compared between day 3 and day 7 of resuscitation. There was no improvement in MODS in the control group, and the difference between means was 0.5893 ± 0.9370 (95% CI -1.434 to 2.613, p=0.5402), whereas, in the centhaquine group, the difference between means was -0.5485 ± 0.3421 (95% CI -1.231 to 0.134, p=0.1135). The change in control was towards worsening (MODS from 1.138 to 1.727), whereas, in centhaquine patients, it was towards improvement (MODS from 1.367 to 0.8182). Centhaquine treatment decreased MODS, whereas control increased and worsened (Figure 9 and Table 4). The number of patients with 2 or more MODS scores on day 7 was significantly less in the centhaquine (13.64%) group than the control (45.45%) group (OR 5.278, 95% CI 0.8522 to 23.32, p=0.0444).

### All-cause Mortality

In the control arm, 28-day all-cause mortality was 11.76% compared to 2.94% in the centhaquine arm (OR: 4.40; 95% CI 0.9651-23.74) with an 8.8% absolute reduction in mortality. Centhaquine treatment led to a numerical decrease in 28-day all-cause mortality in hypovolemic shock patients, statistically insignificant (p=0.0742) at the 95% confidence level but significant at the 90% confidence level.

### Safety and Tolerability

Centhaquine was well tolerated, and a repeat dose, if needed, was administered to the patient without any sequel. No clinically significant effect of the study drug was observed on biochemical or hematological parameters (Table 5). Nine adverse events were reported in nine patients out of 105 patients included in the study. Four adverse events were reported in four control group patients (N=34). All these four events were serious (death, N=4). Five adverse events were reported in five patients of the centhaquine group (N=71). In the centhaquine group, two events in two patients were serious (death, N=2). Three adverse events were reported in three patients from the centhaquine group (Table 6). Two patients had elevated serum creatinine levels (moderate severity), and one patient had vomiting (mild severity), where the outcome resolved without any sequelae with medical intervention. None of these adverse events were related to the drug treatment.

**Table 5.** Haematological, biochemical and serum electrolytes levels of patients

|  | Control (N=34) |  |  | Centhaquine (N=68) |  |  |
| --- | --- | --- | --- | --- | --- | --- |
|  | Day 1<br>(Baseline) | Day 3 | Day 28 | Day 1<br>(Baseline) | Day 3 | Day 28 |
| **Haematology:** |  |  |  |  |  |  |
| *Haemoglobin (g/dL)* | 10.41 ± 0.47 | 10.29 ± 0.42 | 12.37 ± 0.23 | 9.64 ± 0.35 | 9.96 ± 0.30 | 11.56 ± 0.29 |
| *Haematocrit (%)* | 31.90 ± 1.56 | 31.15 ± 1.16 | 37.72 ± 0.85 | 29.48 ± 1.09 | 30.57 ± 0.79 | 36.27 ± 0.74 |
| *Red blood cells (10<sup>3</sup>/mm<sup>3</sup>)* | 3.66 ± 0.17 | --- | 4.32 ± 0.10 | 3.43 ± 0.12 | --- | 4.23 ± 0.09 |
| *White blood cells (/mm<sup>3</sup>)* | 12429.12 ±<br>960.93 | --- | 7322.40 ±<br>320.40 | 12166.04 ±<br>693.06 | --- | 7896.55 ±<br>290.31 |
| *Neutrophils (%)* | 77.83 ± 2.23 | --- | 59.81 ± 1.30 | 77.07 ± 1.47 | --- | 62.81 ± 1.18 |
| *Lymphocytes (%)* | 17.32 ± 1.88 | --- | 32.29 ± 1.24 | 17.28 ± 1.16 | --- | 30.65 ± 1.05 |
| *Monocytes (%)* | 2.77 ± 0.45 | --- | 3.56 ± 0.67 | 2.74 ± 0.27 | --- | 2.93 ± 0.29 |
| *Eosinophils (%)* | 1.97 ± 0.29 | --- | 3.20 ± 0.38 | 2.76 ± 0.39 | --- | 3.65 ± 0.33 |
| *Basophils (%)* | 0.08 ± 0.04 | --- | 0.22 ± 0.08 | 0.10 ± 0.03 | --- | 0.20 ± 0.05 |
| *Reticulocytes (%)* | 1.92 ± 0.21 | --- | 1.28 ± 0.11 | 1.50 ± 0.10 | --- | 1.29 ± 0.07 |
| *Mean corpuscular volume (fL)* | 87.97 ± 1.20 | --- | 87.17 ± 0.95 | 85.75 ± 0.88 | --- | 86.50 ± 0.63 |
| *Mean corpuscular haemoglobin (Pg)* | 28.83 ± 0.52 | --- | 28.79 ± 0.44 | 28.06 ± 0.43 | --- | 28.67 ± 0.27 |
| *Platelets (/mm<sup>3</sup>)* | 192294.12 ±<br>16816.75 | 228160.00 ±<br>14019.59 |  | 188776.12 ±<br>9260.97 |  | 230645.45 ±<br>9755.28 |
| **Lipid profile:** |  |  |  |  |  |  |
| *Triglyceride (mg/dL)* | 112.98 ± 6.04 | --- | 128.87 ± 7.61 | 129.63 ± 6.94 | --- | 136.51 ± 7.55 |
| *Total cholesterol (mg/dL)* | 134.78 ± 7.43 | --- | 149.33 ± 4.98 | 143.43 ± 4.72 | --- | 157.05 ± 4.43 |
| *High-density lipoprotein (mg/dL)* | 39.19 ± 1.79 | --- | 44.43 ± 1.49 | 41.52 ± 1.18 | --- | 43.28 ± 1.06 |
| *Low-density lipoprotein (mg/dL)* | 73.35 ± 5.75 | --- | 80.59 ± 5.35 | 77.41 ± 4.02 | --- | 89.87 ± 3.72 |
| *Very-low-density lipoprotein (mg/dL)* | 24.48 ± 1.56 | --- | 28.69 ± 1.86 | 28.18 ± 1.49 | --- | 29.24 ± 1.53 |
| **Kidney function:** |  |  |  |  |  |  |
| *Serum creatinine (mg/dL)* | 1.28 ± 0.13 | 1.05 ± 0.10 | 0.86 ± 0.04 | 1.22 ± 0.08 | 1.18 ± 0.12 | 0.88 ± 0.03 |
| *Blood urea nitrogen (mg/dL)* | 19.32 ± 3.19 | --- | 12.08 ± 0.51 | 16.28 ± 1.03 | --- | 11.91 ± 0.47 |
| *Glomerular filtration rate*<br>*(ml/min/1.73 m<sup>2</sup>)* | 78.84 ± 5.42 | 93.68 ± 6.60 | 106.22 ± 6.41 | 77.41 ± 4.30 | 84.18 ± 4.04 | 96.55 ± 3.51 |
| **Liver function:** |  |  |  |  |  |  |
| *Alanine aminotransferase (U/L)* | 60.14 ± 26.58 | --- | 23.05 ± 1.60 | 58.73 ± 12.96 | --- | 25.06 ± 2.18 |
| *Aspartate aminotransferase (U/L)* | 121.29 ± 73.15 | --- | 26.66 ± 2.26 | 67.19 ± 10.50 | --- | 24.97 ± 1.92 |
| *Serum bilirubin (mg/dL)* | 0.99 ± 0.07 | --- | 0.72 ± 0.03 | 1.07 ± 0.13 | --- | 0.72 ± 0.03 |
| *Alkaline phosphatase (IU/L)* | 108.45 ± 11.13 | --- | 112.78 ± 12.39 | 121.63 ± 13.35 | --- | 108.41 ± 8.68 |
| *Serum albumin (g/dL)* | 3.54 ± 0.10 | --- | 3.98 ± 0.08 | 3.55 ± 0.07 | --- | 3.87 ± 0.06 |
| **Blood glucose (mg/dL):** | 140.23 ± 9.43 | --- | 95.90 ± 4.32 | 139.06 ± 10.54 | --- | 104.26 ± 4.21 |
| **Serum electrolyte:** |  |  |  |  |  |  |
| *Sodium (mmol/L)* | 136.38 ± 0.96 | --- | 137.23 ± 1.17 | 136.49 ± 0.65 | --- | 137.37 ± 0.63 |
| *Potassium (mmol/L)* | 4.13 ± 0.16 | --- | 3.96 ± 0.05 | 4.16 ± 0.12 | --- | 3.89 ± 0.05 |
| *Calcium (mmol/L)* | 1.78 ± 0.09 | --- | 2.05 ± 0.10 | 1.80 ± 0.07 | --- | 2.06 ± 0.06 |
| **Arterial blood gases** |  |  |  |  |  |  |
| *pH* | 7.32 ± 0.02 | 7.40 ± 0.01 | --- | 7.34 ± 0.01 | 7.40 ± 0.01 | --- |
| *pCO<sub>2</sub> (mmHg)* | 33.65 ± 1.36 | 36.00 ± 1.50 | --- | 31.98 ± 0.90 | 35.49 ± 0.71 | --- |
| *paO<sub>2</sub> (mmHg)* | 117.04 ± 9.39 | 96.01 ± 5.42 | --- | 113.48 ± 5.62 | 99.12 ± 3.70 | --- |
| *FiO<sub>2</sub>* | 0.26 ± 0.02 | 0.22 ± 0.01 | --- | 0.27 ± 0.02 | 0.22 ± 0.01 | --- |

Data are presented as mean ± standard error of the mean. pH- Power of hydrogen; paO<sub>2</sub>- Partial pressure of oxygen; pCO<sub>2</sub>- Partial pressure of carbon dioxide; FiO<sub>2</sub>-Fraction of inspired oxygen.

**Table 6.** Safety of centhaquine and incidence of adverse events

| **Event** | **Control (N=34)** | **Centhaquine (N=68)** |
| --- | --- | --- |
| **Adverse Events of any grade** |  |  |
| Increase in blood Creatinine | 0 (0%) | 2 (2.94%) |
| Vomiting | 0 (0%) | 1 (1.47%) |
| **Serious Adverse Events** |  |  |
| Deaths | 4 (11.76%) | 2 (2.94%) |

## DISCUSSION

Efforts to develop an effective resuscitative agent have not been successful. Although the use of blood products in ratios that epitomize blood transfusion has been observed to improve outcomes [23, 42], nonetheless, fluids and vasopressors are the main elements of resuscitation, they have undesired effects [2, 28, 29]. This randomized multicenter trial results suggest that centhaquine is an effective resuscitative agent for hypovolemic shock. Its resuscitative action is based upon stimulation of venous α_2B_ adrenergic receptors to produce constriction, increase venous return to the heart, increase cardiac output and tissue perfusion [31, 32]. It has no beta-adrenergic activity, mitigating the risk of arrhythmias.

In this trial, the reasons for hypovolemic shock and the extent of blood loss were similar in the control and centhaquine groups. Blood pressure and lactate levels at the time of enrollment were similar in both groups. In India and neighboring countries, the standard of care in critically ill patients involves fluid therapy keeping in mind cumulative fluid balance and the use of vasopressors [43]. Such an approach is common across the globe and was followed in the present study. During the first 48 hours of resuscitation, fluids, blood, blood products, vasopressors, and urine output was similar in both groups. A significant number of patients with improved SBP and DBP were in the centhaquine group than the control group. A 7.44% decrease in AUC of the mean difference in DBP from baseline in the centhaquine group than the control group allows more ventricular filling, increasing cardiac output. An increase in AUC of 12.99% in SBP of the centhaquine group than the control group indicates that the rate of venous return to the heart is higher in the centhaquine group than the control group. The primary determinant of pulse pressure is the stroke volume, and a 48.14% increase in AUC of the mean difference in pulse pressure from baseline in the centhaquine group than the control group indicates a significant increase in stroke volume due to centhaquine.

Hypovolemic shock results in a drop in cardiac output, lowering tissue and organ blood perfusion, leading to multiple organ failure and death. Vasopressors increase blood pressure by arterial constriction and increasing heart rate. Cardiac output can increase due to an increase in heart rate, but it does not account for the total increase in cardiac output [44]. Sixty to seventy percent of the total blood volume pooled in the venous system is adjustable [45]. The venous system is critically important following hemorrhage because it can be used to mobilize pooled (unstressed) blood volume towards systemic (stressed) circulation [46, 47]. In a patient with hypovolemic shock, centhaquine converts the venous unstressed blood volume to stressed blood volume and improves cardiac output and blood circulation, making it an ideal candidate to resuscitate patients.

Shock index (SI), defined as heart rate divided by SBP, has a normal range of 0.5 to 0.7 in healthy subjects. SI ≥ 1.0 has been associated with significantly poorer outcomes in patients with acute circulatory failure [48, 49]. SI also indicated a rapid improvement of left ventricular function in centhaquine compared to the control group. SI, an important prognostic indicator, is linearly inversely related to physiologic parameters, such as cardiac index, stroke volume, left ventricular stroke work, and mean arterial pressure [48, 50, 51]. SI significantly improved (P<0.0001) in the centhaquine group in the first 4 hours of resuscitation. A difference between centhaquine and control groups was observed within the first hour of resuscitation, where a decrease in SI was significant (0.1505 ± 0.0682; p=0.0321) in centhaquine compared to the control group. The initial hours of resuscitation are the most critical in improving the outcome of these patients.

These findings support our preclinical studies in rat, rabbit, and swine models of hemorrhagic shock, where centhaquine significantly decreased blood lactate and increased mean arterial pressure, pulse pressure, and cardiac output [34–36, 52]. The safety and efficacy of centhaquine were evaluated in healthy volunteers and hypovolemic shock patients in phase I (CTRI/2014/06/004647; NCT02408731) and phase II studies. In the phase II study (CTRI/2017/03/008184; NCT04056065), centhaquine demonstrated significant efficacy in the improvement of vital clinical parameters [39, 40].

Under conditions of shock, inadequate blood flow to the tissues results in increased blood lactate levels. High blood lactate levels and an increase in the base deficit in patients are suggestive of poor outcomes and high mortality rates [53]. Early lactate clearance is associated with a decrease in mortality, shorter ICU length of stay, and duration of mechanical ventilation [54]. In the present study, the number of patients on day 3 of resuscitation having a blood lactate level of 1.5 mmol/L or less was 46.88% in the control group and 69.35% in the centhaquine group (p=0.0336). Similarly, the base deficit of less than -2 mmol/L on day 3 of resuscitation was significantly (p=0.0137) more in the centhaquine group (69.84%) than in the control group (43.75%).

Baseline GCS scores were similar in both control and centhaquine groups (Table 4), indicating a similarity in the neurological status between the two groups. Centhaquine significantly improved ARDS and MODS. Studies in a swine model of hemorrhagic shock showed that centhaquine significantly improved Horowitz index (327 ± 10 and 392 ± 16 in the control and centhaquine group, respectively) and reduced pulmonary edema [31, 36]. This study indicates an improvement in the ARDS of patients with hypovolemic shock. In the control group, there was an improvement in ARDS on day 3 compared to day 1, but it was not statistically significant. Whereas in the centhaquine group, a significant (p=0.0449) improvement in ARDS was observed on day 3 of resuscitation compared to day1. MODS in the control group increased on day 7 compared to day 3, while in the centhaquine group, it decreased on day 7 compared to day 1. A direct comparison of MODS on day 7 between the control and centhaquine group revealed that MODS of less than 2 was in 86.36% of centhaquine group patients compared to 54.55% patients in the control group (p=0.0444).

It was recognized that 55% of all trauma patients have hypocalcemia [55], which worsens by infusion of blood and blood products having citrate (used for storage), which chelates calcium when infused. A drop in calcium can aggravate coagulopathy leading to continued hemorrhage [56]. Calcium levels in the present study were similarly improved from 1.78 ± 0.09 to 2.05 ± 0.10 mmol/L in the control group and from 1.80 ± 0.07 to 2.06 ± 0.06 mmol/L in the centhaquine group indicating that centhaquine did not affect serum calcium. Centhaquine did not affect serum sodium and potassium levels and other biochemical parameters (Table 5).

An improvement in all the above clinical and biological markers appears to contribute towards improved outcomes and reduced deaths in the centhaquine group. Mortality is the primary outcome for most clinical trials in critical care medicine; however, many factors can influence the outcome [57]. In a meta-analysis of trials with study intervention, a reduction in mortality was reported in 27 randomized controlled trials of 15,612 patients and increased mortality in 16 randomized controlled trials of 10,462 patients [57]. These trials were carried out in the general ICU population or patients with sepsis, while no specific study was reported in hypovolemic shock patients. Upon further analysis, only 13 randomized controlled trials demonstrated reduced mortality rates, which were attributed to disease conditions rather than demonstrating an effect of a new therapy [57].

In summary, none of the new therapies has demonstrated any reduction in mortality. We have carefully included a patient population that was more homogeneous to avoid heterogeneous factors influencing the outcome to determine the effect of the intervention (centhaquine) on clinical outcome. To our knowledge, this is the only late-stage clinical study that has demonstrated a significant survival advantage with an 8.8% absolute reduction in mortality. Centhaquine was safe and well-tolerated with no drug-related adverse events. Centhaquine obtained Marketing Authorization from the regulatory authorities in India to treat hypovolemic shock patients in 2020. We did a meta-analysis of mortality data we obtained from phase II and III studies because the inclusion criteria were similar and found that mortality in the control group (N=56) was 10.71%, and in the centhaquine group (N=91) it was 2.20% (OR 5.340, 95% CI 1.27 to 26.50, p=0271) which is statistically significant at 95% confidence interval.

Mortality observed in our study in the control group is a little lower related to previous studies. In a study that analyzed 4038 patients from 120 ICUs in India, a mortality rate was 20.8% in well-equipped ICUs [58]. Further analysis showed that in trauma patients, mortality was 14.1% (26 out of 185) [58] which is slightly higher than that observed in the control group (11.8%) of our study. Similarly, the Australia India Trauma Systems Collaboration registry data of 9354 patients showed 30-day mortality of 12.4% [59]. Treatment with centhaquine produced an absolute reduction in 28-day all-cause mortality by 8.8%.

The effect of centhaquine on systemic hemodynamics of patients with hypovolemic shock depends on the fluid status. A limitation of this study is that we have not examined the effect of centhaquine on the volume status of patients with hypovolemic shock. Centhaquine was administered in a total volume of 100 mL over 60 minutes; this is a small volume and not likely to cause any volume overload. Moreover, the total volume of fluids administered in control and centhaquine groups was similar in the first 48 hours. Blood products administered in the first 48 hours of resuscitation were similar in control and centhaquine groups. In the first 48 hours, the urine output was not different in control and centhaquine groups. Another limitation of this study is that although it is a multicenter study, it was conducted exclusively in patients from India. We recognize that the demographics and SOC may vary in other countries. In this study, many patients missed the Golden Hour resulting in a greater possibility of developing secondary complications. Delayed intervention is likely to cause the release of inflammatory and apoptotic substances, producing additional organ damage and failure of multiple organs resulting in higher mortality. In countries where the patients are likely to be resuscitated within the Golden Hour, secondary complications are less likely, and we expect greater effectiveness of centhaquine.

The therapeutic potential of centhaquine for other forms of shock associated with hemodynamic instability or refractory hypotension resulting in multi-organ failure and ultimately death may be explored in future studies. Some of these conditions may include septic shock, and in fact, a significant number of COVID-19 patients die from shock with or without sepsis [60, 61]. There are few drug candidates under development for sepsis with the aim of reducing organ dysfunction [62]. Septic shock is a type of distributive shock where a significant shift occurs within the vascular compartment and out of the vascular system resulting in a state of hypovolemia managed by administration of fluids and vasopressors [63]. Centhaquine can be helpful in septic shock management like hypovolemic shock by augmenting cardiac output and improving tissue blood perfusion. Other investigators and we are likely to initiate studies to determine the efficacy of centhaquine in septic shock patients.

How does centhaquine fit in a typical resuscitation protocol? Patients with uncontrolled bleeding undergo damage-control resuscitation to stop the blood loss and initiate resuscitation, keeping in mind permissive hypotension targeting mean arterial pressure at 65 mmHg [64, 65]. Resuscitation with centhaquine is likely to limit the use of vasopressors and may help achieve vasopressor-free resuscitation [46]. If required, a balanced resuscitation follows by transfusing blood or blood products in a ratio similar to whole blood [64].

### Conclusion

Centhaquine (Lyfaquin^®^) is a highly effective resuscitative agent for treating hypovolemic shock as an adjuvant to SOC.

## Data Availability

The data are not publicly available due to restrictions that could compromise the privacy of research participants. The anonymized patient datasets generated during and/or analyzed during the current study are available from the corresponding author on reasonable request from a bona fide researcher/research group including
Supplemental Tables 2 and 3.
The study protocol (PMZ-2010/CT-3.1/2018) was approved by the Drug Controller General of India (DCGI), Ministry of Health and Family Welfare, Government of India, and Institutional Ethics Committee of all the 14 Institutions.

## DISCLOSURES

### Funding

Pharmazz, Inc. Willowbrook, IL, USA, provided funding for this study and paid the open access fee.

### Conflict

Anil Gulati has issued and pending patents and is an employee and stockholder of Pharmazz, Inc. Rajat Choudhuri, Ajay Gupta, Saurabh Singh, S.K. Noushad Ali, Gursaran Kaur Sidhu, Parvez David Haque, Prashant Rahate, Aditya R. Bothra, Gyan P. Singh, Sanjeev Maheshwari, Deepak Jeswani, Sameer Haveri, Apurva Agarwal, and Nilesh Radheshyam Agrawal declare no conflicts of interest that are directly relevant to the content of this article.

### Ethics approval

The study protocol (PMZ-2010/CT-3.1/2018) dated July 16, 2018, was approved by the Drugs Controller General of India (DCGI), Directorate General of Health Services, Ministry of Health & Family Welfare, Government of India (DCGI CT NOC. No.: CT/ND/66/2018). Furthermore, each institutional ethics committee reviewed and approved the study protocol before initiating patient enrolment.

### Consent to participate

Written informed consent was obtained from all patients or their legally authorized representatives.

### Consent for publication

Not applicable

### Code availability

Not applicable

### Data and material

The anonymized patient datasets generated during and/or analyzed during the current study are available from the corresponding author on reasonable request from a bona fide researcher/research group.

### Authors’ contributions

Study concept and design: AG; Investigation: RC, AG, SS, SKNA, GKS, PDH, PR, ARB, GPS, SM, DJ, SH, AA, and NRA; Acquisition of data: RC, AG, SS, SKNA, GKS, PDH, PR, ARB, GPS, SM, DJ, SH, AA, and NRA; Analysis and interpretation of data: AG; Drafting of the manuscript: AG; Review of the manuscript: RC, AG, SS, SKNA, GKS, PDH, PR, ARB, GPS, SM, DJ, SH, AA, and NRA; Funding acquisition: AG.

## Acknowledgments

We thank all the participants, patients, and their families for the study.

## Notes

### Competing Interest Statement

Anil Gulati is a stockholder and employee of Pharmazz, Inc. All other authors have no competing interest to declare.

### Clinical Trial

NCT04045327 (Clinical Trials Registry, India CTRI/2019/01/017196). The study protocol (PMZ-2010/CT-3.1/2018) was approved by the Drug Controller General of India (DCGI), Ministry of Health and Family Welfare, Government of India and Institutional Ethics Committee of all the 14 Institutions.

### Funding Statement

The present study was funded by Pharmazz, Inc. Toxicological study of centhaquine in dog was supported by the National Heart, Lung, and Blood Institute of the National Institutes of Health under Award Number R43HL137469.

### Summary of Updates

The anonymized patient datasets generated during and/or analyzed during the current study are available from the corresponding author on reasonable request from a bona fide researcher/research group including Supplemental Tables 2 and 3. This version of the manuscript has been submitted to the journal Drugs.

