## Supplemental Table 1 for "A multicentric, randomized, controlled phase III study of centhaquine (Lyfaquin^®^) as a resuscitative agent in hypovolemic shock patients"

**Supplementary Table 1** List of participating institutions and hospitals

| **S. No.** | **Study site** | **Enrolled patients** |
| --- | --- | --- |
| 1 | Institute of Post Graduate Medical Education and Research and SSKM Hospital, 244 A.J.C. Bose Road, Kolkata 700020, West Bengal | 25 |
| 2 | King George’s Medical University, Lucknow 226003, Uttar Pradesh | 04 |
| 3 | Institute of Medical Sciences, Banaras Hindu University, Varanasi 221005, Uttar Pradesh | 11 |
| 4 | Seven Star Hospital, Jagnade Square, KDK College Road, Nagpur 440009, Maharashtra | 06 |
| 5 | Rahate Surgical Hospital, 517 Juni Mangalwadi, Central Avenue, Nagpur 440008, Maharashtra | 05 |
| 6 | New Era Hospital and Research Institute, Queta Colony, Central Avenue Road, Nagpur 440008, Maharashtra | 01 |
| 7 | Sidhu Hospital Pvt. Ltd. G.T. Road, Doraha 141421, Ludhiana, Punjab | 09 |
| 8 | Ganesh Shankar Vidyarthi Memorial Medical College, Swaroop Nagar, Kanpur 208002, Uttar Pradesh | 02 |
| 9 | KLE’s Dr. Prabhakar Kore Hospital and Medical Research Centre, Nehru Nagar, Belgaum 590010, Karnataka | 02 |
| 10 | Criticare Hospital and Research Institute, Dhanshree Complex, Sitabuldi, Nagpur 440012, Maharashtra | 02 |
| 11 | Christian Medical College and Hospital, Brown Road, Ludhiana 141008, Punjab | 06 |
| 12 | Jawahar Lal Nehru Medical College and Attached Hospitals, Kala Bagh, Ajmer 305001, Rajasthan | 02 |
| 13 | AC Subba Reddy Government Medical College and Hospital, Dargamitta, G.T. Road, Nellore 524004, Andhra Pradesh | 10 |
| 14 | Chiranjeev Medical Centre, Shivaji Nagar, Jhansi 284128, Uttar Pradesh | 20 |
|  | **Total** | **105** |
